# A role for Nucleocapsid-specific antibody function in Covid-19 Convalescent plasma therapy

**DOI:** 10.1101/2022.02.19.22271230

**Authors:** Jonathan D. Herman, Chuangqi Wang, John Stephen Burke, Yonatan Zur, Hacheming Compere, Jaewon Kang, Ryan Macvicar, Sally Shin, Ian Frank, Don Siegel, Pablo Tebas, Grace H. Choi, Pamela A. Shaw, Hyunah Yoon, Liise-anne Pirofski, Boris Juelg, Katharine J. Bar, Douglas Lauffenburger, Galit Alter

## Abstract

COVID-19 convalescent plasma (CCP), a passive polyclonal antibody therapeutic, has exhibited mixed results in the treatment of COVID-19. Given that the therapeutic effect of CCP may extend beyond the ability of SARS-CoV-2-specific antibody binding and neutralization to influence the evolution of the endogenous antibody response, we took a systematic and comprehensive approach to analyze SARS-CoV-2 functional antibody profiles of participants in a randomized controlled trial of CCP treatment of individuals hospitalized with COVID-19 pneumonia where CCP was associated with both decreased mortality and improved clinical severity. Using systems serology, we found that the clinical benefit of CCP is related to a shift towards reduced inflammatory Spike (S) responses and enhanced Nucleocapsid (N) humoral responses. We found CCP had the greatest clinical benefit in participants with low pre-existing anti-SARS-CoV-2 antibody function, rather than S or N antibody levels or participant demographic features. Further, CCP induced immunomodulatory changes to recipient humoral profiles persisted for at least two months, marked by the selective evolution of anti-inflammatory Fc-glycan profiles and persistently expanded nucleocapsid-specific humoral immunity following CCP therapy. Together, our findings identify a novel mechanism of action of CCP, suggest optimal patient characteristics for CCP treatment, identify long-last immunomodulatory effects of CCP, and provide guidance for development of novel N-focused antibody therapeutics for severe COVID-19 hyperinflammation.

## Introduction

The COVID-19 pandemic has claimed more than 4.5 million lives to date.(Dong et al., 2020) Despite the development and deployment of vaccines to prevent severe COVID-19 and hospitalization, a significant portion of the world remains unvaccinated. Moreover, the evolution of SARS-CoV-2 variants of concern that are more infectious and able to evade both vaccine and monoclonal therapeutic immunity fuels an urgent need for effective therapeutics for hospitalized individuals with severe COVID-19.

Due to its immediate availability, use, and safety in prior respiratory pandemics, COVID-19 convalescent plasma (CCP) was proposed as a modality to treat COVID-19 from the start of the pandemic.(Casadevall and Pirofski, 2020) However, evidence of CCP clinical efficacy has been mixed. Smaller randomized clinical trials and observational matched case-control studies have shown a benefit of high-titer CCP in patients early in the course of COVID-19.(Avendaño-Solá et al., 2021; Joyner et al., 2021; Libster et al., 2021; O’Donnell et al., 2021; Salazar et al., 2021; Sullivan et al., 2021; Yoon et al., 2021) However, larger trials have not found an overall benefit of CCP, with the caveat that many of these trials treated patients with severe COVID-19 at later stages of disease.(Group et al.; Investigators et al., 2021) Along these lines, the CONCOR-1 trial, a large, randomized controlled trial of CCP in hospitalized COVID-19 patients, did not find a clinical benefit of CCP and found that low-titer CCP may have been harmful in severely ill patients.(Bégin et al., 2021) However, deeper immunologic analysis of the CCP used in CONCOR-1 found that antibody-dependent cell cytotoxicity (ADCC) was associated with a lower risk of intubation or death by day 30,(Bégin et al., 2021) suggesting that the efficacy of CCP may in part depend on antibody Fc-effector functions. A possible role for antibodies with Fc-functional activity highlights the fact that antibodies can leverage the immune system to attenuate disease via a broad array of effector functions. The importance of these functions in CCP therapy have not been fully elucidated.

Previous studies have highlighted the remarkable heterogeneity of SARS-CoV-2 specific humoral immune responses across CCP products, marked by different SARS-CoV-2 antibody titers and antibody-effector functions.(Herman et al., 2021b) However, whether particular functions or antibody qualities, including ADCC, are associated with differential therapeutic outcome remains incompletely understood. Thus, we applied system serology to an open-label randomized clinical trial that showed evidence of a mortality benefit from CCP treatment with Receptor-binding domain (RBD) ELISA-selected CCP treatment.(Bar et al., 2021) We found that CCP treatment delayed the evolution of Spike (S)-specific inflammatory antibody responses and induced stronger N-specific antibody responses, both of which were associated with improved outcomes in CCP-treated patients. While pre-existing SARS-CoV-2 specific humoral responses were inversely correlated with response to CCP therapy, participants with the lower antibody function rather than low antibody levels experienced the strongest clinical benefit from CCP. Further, we not only found that CCP modulated humoral immunity during acute disease, but also two months after treatment leading to more anti-inflammatory S-specific Fc glycans and persistent N-specific immunodominance.

## Results

### Global SARS-CoV-2 humoral profiles of CCP-treated and control participants

With the emergence of novel SARS-CoV-2 variants that can escape both vaccine-induced neutralizing antibody responses and monoclonal antibody therapeutics, CCP has regained attention as a potential therapeutic strategy to treat COVID-19.(Focosi et al., 2021; O’Connell et al., 2021; Schmidt et al., 2021) However, the results of clinical trials with CCP have been mixed, related to the striking heterogeneity of CCP, and our incomplete understanding of the mechanisms of action of this natural therapeutic agent.(Bégin et al., 2021; Herman et al., 2021b; Morgenlander et al., 2021; Natarajan et al., 2021; Wang et al., 2020) To gain a more granular understanding of the CCP properties that contribute to therapeutic efficacy, we profiled the SARS-CoV2-specific antibody response across a group of patients enrolled in a randomized control trial of COVID-19 convalescent plasma (CCP) conducted at University of Pennsylvania.(Bar et al., 2021) The UPenn CCP2 trial enrolled 80 individuals hospitalized with COVID-19 pneumonia (defined as a positive SARS-CoV-2 PCR assay, SaO2 < 93% on room air or supplement oxygen use, and radiologic evidence of pneumonia) as demonstrated in Figure 1A. One participant declined CCP administration and withdrew from the study early, thus we included 79 participants in our analyses; 40 of whom were randomized to receive two units of CCP plus standard of care and 39 received standard of care alone. Participants median age was 63 years (IQR [52, 74]), 54% were female, 13% were on immunomodulatory treatments at baseline, 26% were diagnosed with cancer, and prior to CCP randomization 81% of participants were treated with remdesivir and 83% of participants were treated with corticosteroids.

**Figure 1:**
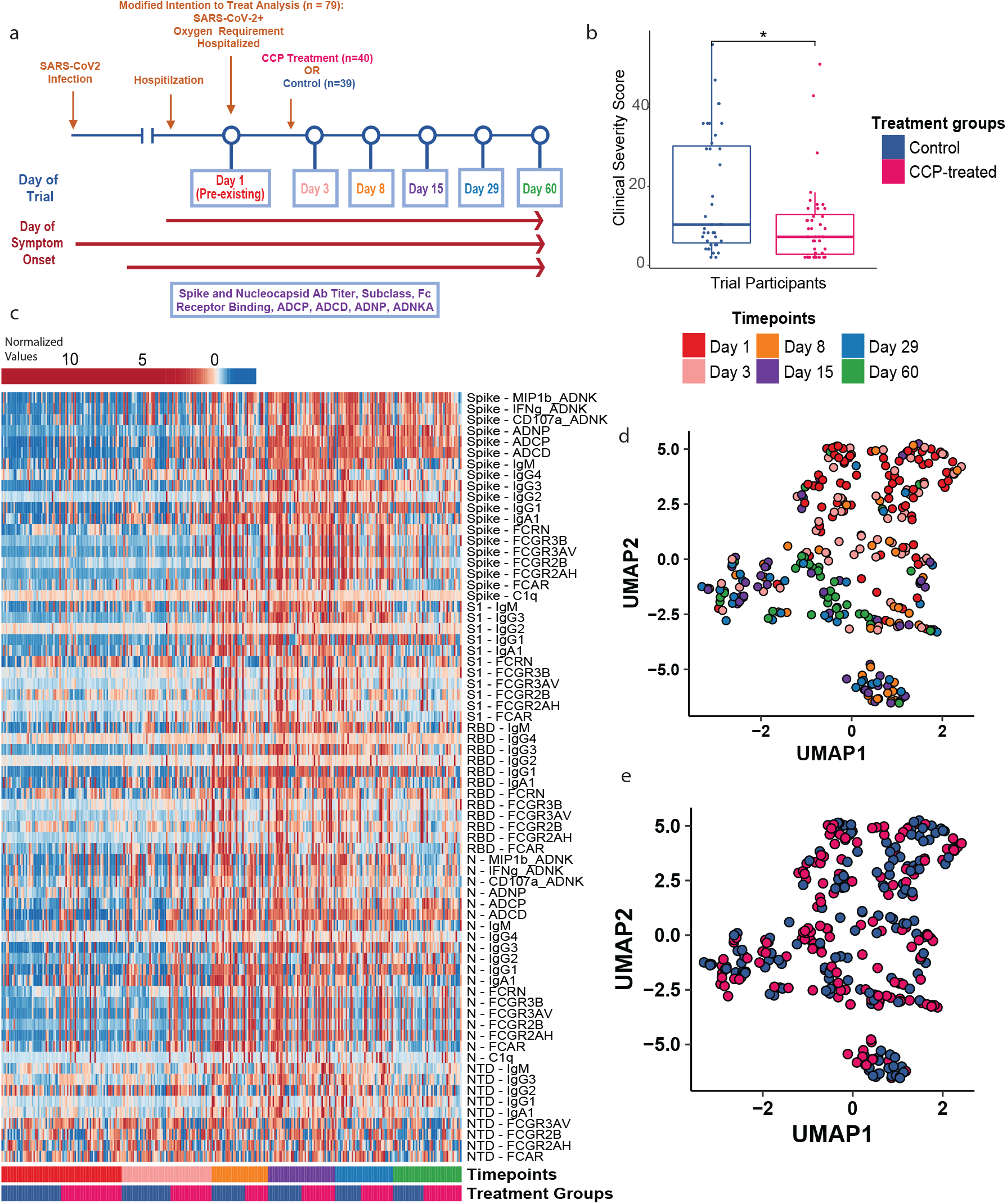
Global Anti-SARS-CoV2 Response in CCP-treated and control individuals. (a) Schematic of the UPenn CCP2 Randomized Clinical trial of COVID-19 Convalescent Plasma and the antibody profiling performed in this paper. In total, we profiled 302 samples from 79 patients. Patients were randomly assigned to CP treatment (n=40) or Standard of Care (n=39). Patient serum samples were collected at day 1 (n=79), day 3 (n=59), day 8 (n=37), day 15 (n=44), day 29 (n=38), and day 60 (n=45). Since patients experienced symptomatic COVID-19 for variable number of days prior to presenting to the hospital, we have organized patient serum samples by Day of the Trial (Day = 1 enrollment in clinical trial) and by Day of Symptom onset (Day 1 = first day of COVID-19 associated symptoms). (b) Clinical Severity Score in CCP-treated and control groups. Significance corresponds to a two-sided Wilcoxon test p-values (p value = 0.0333) (*: p < 0.05) (c) Heatmap of the SARS-CoV-2-specific antibody profiles of all patient time points, arranged by time point, arm of the trial, and patient age. (d-e) Uniform Manifold Approximation and Projection (UMAP) was used to visualize the multivariate SARS-CoV-2 antibody profiles in two dimensions. Each point represents a given patient timepoint and colors indicate (d) time point of collection and (e) treatment group.

The CCP2 trial enrolled participants early in their disease course, with a median 6 days of symptoms and 1 day of hospitalization. Mortality and the Clinical Severity Score (CSC) were the two prespecified outcomes of the this trial.(Bar et al., 2021) The Clinical Severity Score (CSC) is a composite score that aims to effectively rank patients based on their disease severity, taking into account multiple endpoints in a prioritized manner, as described by Shaw and Fay 2016.(Shaw and Fay, 2016) The CSC was determined by a participant’s survival time, time to recovery, and disease course while in the hospital including the 8-point WHO ordinal score (WHO8), use of supplemental oxygen and adverse events. The CSC was found to be significantly different between CCP recipients and controls (median [IQR] 7 [2.75, 12.5] vs. 10[5.5, 30]; p=0.037 by Wilcoxon rank sum test) (Figure 1B).(Bar et al., 2021) Additionally, the study also found a mortality benefit associated with CCP administration at day 28 (OR 0.156, p=0.013), with 5% (2 of 40) vs. 25.6% (10 of 39) mortality in CCP-treated vs. control participants. These clinically meaningful outcomes provided an opportunity to comprehensively examine the immunological profiles across CCP-treated and untreated individuals to define potential biomarkers of immunity.

SARS-CoV-2-specific responses were deeply profiled across CCP-treated and control participants using Systems Serology.(Chung et al., 2015; Herman et al., 2021b) Antigen-specific isotype (IgM, IgA), subclass (IgG1, IgG2, IgG3, and IgG4), and Fc-receptor binding (FcγR2AH, FcγR2B, FcγR3AV, FcγR3B, FcAR, and FcRn) analysis was performed against Spike (S), the S1 domain of S, the receptor binding domain (RBD) of S, the N-terminal domain (NTD) of S, and Nucleocapsid (N) across plasma samples from CCP recipients and control patients at enrollment of the study (pre-CCP or Day 1 mentioned later), at Day 3, Day 8, Day 15, Day 29, and Day 60 after enrollment (Figure 1A). Additionally, antibody-directed innate immune cell functional analysis was performed over time against S and N, including antibody-dependent complement deposition (ADCD), antibody-dependent cell phagocytosis (ADCP), antibody-dependent neutrophil phagocytosis (ADNP), and antibody-dependent NK cell activation (ADNK) (Figure 1C).

As expected, the humoral immune response to SARS-CoV-2 evolved across all the participants (Figure 1C, D). Nearly all S- and N-specific antibody features increased in the first two weeks of SARS-CoV-2 infection (Figure 1C). Multivariate Uniform Manifold Approximation and Projection (UMAP) visualization highlighted the similarities of the two groups’ profiles at the start of the study, with most Day 1 samples (Red) at the top of Figure 1D and most Day 60 samples (Green) at the bottom. These data confirm significant changes in the humoral immune responses over time. Additionally, samples from the CCP-treated and control arms of the study were intermixed throughout the UMAP visualization (Figure 1E). Thus, we proceeded to perform a more detailed analysis to identify whether the evolution of the SARS-CoV-2 specific humoral immune response differed between CCP-treated and control participants.

### Convalescent plasma results in a delay in the development of SARS-CoV-2 anti-Spike inflammatory antibody profiles

Given the similarity in pre-existing (Day 1) anti-SARS-CoV-2-specific antibody profiles across CCP-treated and control participants using UMAP (Figure 2A), Local Inverse Simpson’s Index (LISI) score analysis (Supplementary Figure 1A), and univariate differences (Figure 2B and Supplementary Figure 1B), we examined differences in the trajectories of the humoral immune response across the two groups. We focused on the early evolutionary differences across the groups, over the first two weeks of the trial (Day 1 to 15 of the clinical trial). When we looked at the distribution of how long patients had symptomatic COVID-19 prior to enrollment, we found it varied greatly from one to twenty days (Supplementary Figure 1C). Thus, to adjust for heterogeneity in each participant’s time from COVID-19 symptom onset, all participant humoral profile data were arranged by the days from the onset of symptoms prior to randomization. By week 3 after symptom onset, CCP-treated individuals had lower S-specific titers, Fc-receptor binding, and Ab-dependent functional activity (Figure 2C). This delay in the evolution of the S-specific response was also evident at Day 8, when data was analyzed agnostic of days of symptom onset (Supplementary Figure 1D).

**Figure 2:**
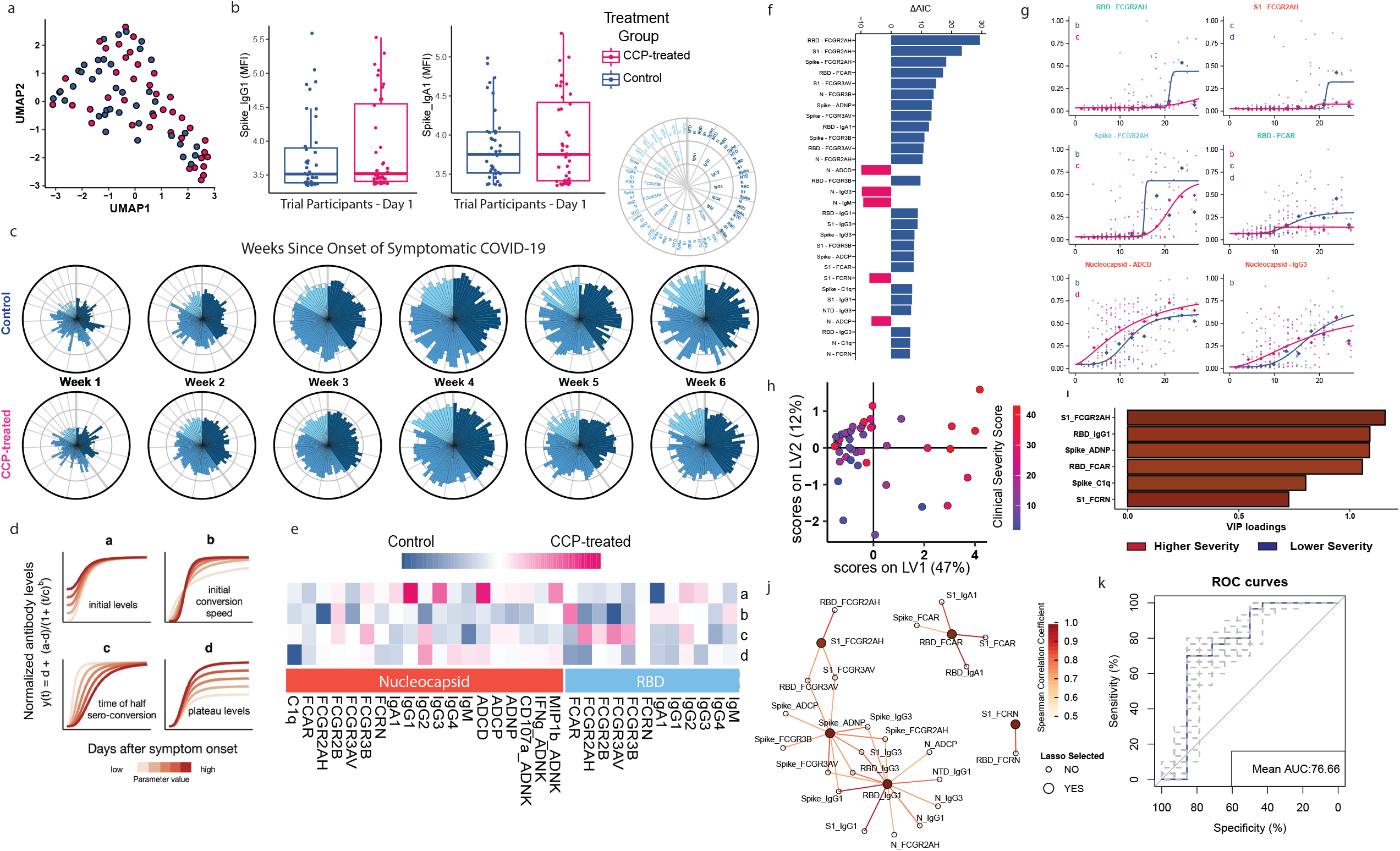
CCP provides clinical benefit by limiting the development of the inflammatory spike antibody trajectory. (a-b) Investigation of pre-existing (Day 1) Spike antibody profiles. (a) UMAP visualization of the samples at Day 1(pre-existing) and (b) univariate box plots on Spike IgG1 and Spike IgA1 of CCP-treated and control patients on Day 1. Each box represents the median (central line) and IQR (25% and 75% percentiles) and the two whiskers represent 1.5 * IQR. (c) The polar plots depict the mean percentile of each antibody features at each week since onset of symptoms across the control arm (top) and CCP-treated arm (bottom). Number of patient samples included per time point as follows Week (# Control, # CCP-treated): Week 1 (15, 15), Week 2 (31, 32), Week 3 (24, 20), Week 4 (14, 15), Week 5 (7, 10), and Week 6 (10, 12). (d-f) We employed a four-parameter logistic regression to fit the antibody growth trajectories to dissect the temporal-specific differences between CCP-treated and control patients for each antibody feature. (d) A visual representation of the logistic regression model and the effect of each parameter on the curve of the model. (e) This heatmap shows the Akaike weighted average parameter differences between the two groups. Each column shows a parameter, which is normalized across the features. The color intensity indicates whether the parameter is higher in the CCP-treated (blue) or control (orange) model. (f). The bar plot depicts the delta-AIC of the best model compared with the model without differences. The higher the delta-AIC, the better the model can explain the trajectory difference. The sign of delta-AIC represents the AUC difference between the CCP-treated and control curves, thereby showing whether the antibody feature is enriched in the CCP-treated model (negative) or the control model (positive). The bars are colored according to whether the feature was enriched in the CCP-treated model (pink) or control model (blue). (g) Plots of the CCP-treated (blue) and control (pink) patient models for selected features that differed most between the groups based on deltaAIC and delta AUC. Each point represents the normalized antibody feature of a single patient timepoint and the line represents the logistic regression model. (h-k) PLS-R regression model that predicts clinical severity in both CCP-treated and control patients based on the top30 features suggested by the four-parameter logistic model. The PLS-R model uses SARS-CoV-2 humoral profiles from Week 3 since the onset of Symptomatic COVID-19. (h) The score plot of PLS-R regression shows the separation of Week 3 samples along the continuum of clinical severity. Each dot represents a patient. (i) The bar graph shows the variable importance in projection (VIP) score in the PLS-R model of the LASSO-selected features. Features are colored by disease severity; features enriched in patients with higher severity (CSC > 20, Red) or lower severity (CSC <= 20, Blue) COVID-19. (j) The network diagram illustrates the co-correlated features that are significantly correlated with the LASSO-selected features (larger nodes) (p-value < 0.05 after multiple test correction using the Benjamini-Hochberg procedure, spearman correlation coefficient > 0.7). Nodes enriched in patients with higher severity or lower severity COVID-19 were colored with red and blue respectively. (k) The ROC curve represents the predictive ability of the Lasso-selected Spike features (S1 FcR2AH, RBD IgG1, Spike ADNP, RBD FcAR, Spike C1q, and S1 FcRn) to distinguish higher severity score (CSC > 20) from low severity score (CSC <= 20) using the PLS-DA model. Five-fold cross-validations were run for 100 times, achieving a mean area under the curve (AUC) of 76.6. The blue line represents the mean ROC curve while the dotted lines represent individual cross-validation ROC curves.

To gain granular insights into the specific humoral immune responses that evolved differentially across the two groups, four-parameter logistic regression models were generated for each antibody feature across each group from Day 0 to Day 28 from symptom onset.(Zohar et al., 2020) This approach allowed us to quantitatively define the parameters that were differentially modulated by CCP treatment by identifying differences in (a) initial levels, (b) initial seroconversion speed, (c) seroconversion time, or (d) final plateau levels (Figure 2D). Though initial levels and initial conversion speeds were mixed for both RBD and N features, final RBD-specific titers (d - plateau level) and FcR-binding (FcγR2a, FcαR, and FcγR3b) were largely higher in the control population (Figure 2E, Supplementary Figures 1F, 2). In contrast, N-specific titers and antibody features exhibited the same plateau levels in both groups or were slightly higher, specifically N-specific IgM, IgG2, and FCγR3b binding levels, in the CCP-treated population (Figure 2E, Supplementary Figure 1F). These data suggested that CCP treatment was associated with a blunting of the evolution of inflammatory anti-S-specific humoral immune profiles in a distinct manner separate from N-specific humoral immunity.

To stratify individual humoral characteristics that differed most across the CCP-treated and control groups, we used the Akaike Information Criterion (AIC) of the paired models to quantify longitudinal differences across groups (Figure 2F, Supplementary Figure 1E). We found that S-, RBD-, and S1-specific FcγR2a binding differed most between the two treatment arms (Figure 2F). Moreover, CCP-treated individuals exhibited lower levels and delayed evolution of S-specific FcγR2a binding antibodies (Figure 2G). Conversely, N-specific ADCD, IgG3, and IgM differed between the two models and were enhanced in CCP-treated individuals (Figure 2F, G). Specifically, N-specific IgG3 and N-specific ADCD developed earlier in CCP-treated individuals and N-specific ADCD reached higher levels in CCP-treated individuals (Figure 2G). Collectively, using a population-based logistic regression model, we found strong evidence that CCP treatment resulted in attenuated inflammatory anti-S immune evolution and was linked to selectively enhanced N-specific humoral immune features.

### CCP-induced blunting of Spike-specific inflammatory antibody features associated with improved clinical outcomes

Given the differences observed in S- and N-specific humoral immune evolution between the CCP-treated and control groups, we next sought to understand whether antibody properties enriched and depleted in CCP-treated participants were associated with improved clinical outcomes (measured by CSC) in both the CCP-treated and control participants.(Bar et al., 2021) Specifically, we selected the 30 antibody features with the greatest |ΔAIC| values from the logistic regression model of Day 0 to Day 28 from symptom onset (Figure 2F). A least absolute shrinkage and selection operator (LASSO) was then applied to identify the minimal features that differed most across the CSC score at week 3 following symptom onset, and a partial least squares regression (PLS-R) was applied to evaluate the association between CSC and the set of LASSO selected features (Figure 2H). Importantly, the PLS-R model identified differences between the groups that were statistically significant (Supplementary Figure 1G and 1H). Only 6 of the top 30 AIC selected features were sufficient to separate all participants based on CSC scores, including S1-specific FcγR2a binding antibody levels, RBD-specific IgG1 levels, S-specific ADNP, RBD-specific FcαR binding levels, S-specific C1q binding levels, and S1-specific FcRn binding. Interestingly, these 6 features were both enriched in controls (Figure 2F) and in those with the most severe disease, defined as participants with a CSC > 20 (Figure 2I).

To discern the antibody properties associated with treatment benefit, we next investigated the associations of LASSO-selected antibody features marking poor clinical trajectories using co-correlation networks. A large co-correlation network connected three of the LASSO-selected features: S1-specific FcγR2AH, S ADNP, RBD IgG1 (Fig 2J). This co-correlation network contained a broad and highly inflammatory S/RBD/S1-specific humoral profile, including more functional antibody subclasses (IgG1 and IgG3), S-specific neutrophil activity (ADNP), and S-specific monocyte responses (FcR2A, ADCP). A second tight co-correlation network linked RBD-specific FcαR binding levels with S-, S1-, and RBD-specific IgA/FcαR features, confirming prior observations that RBD/S-specific IgA responses are associated with worse disease severity.(Bartsch et al., 2021; Ma et al., 2020; Yu et al., 2020; Zervou et al., 2021) A third network consisted of RBD and S1-specific binding to the neonatal Fc-receptor (FcRn). These three co-correlation networks consistently highlight the expanded and highly inflammatory S-specific humoral immune responses in individuals with the greatest disease severity (highest CSC).

To determine if features associated with poor clinical outcomes in the non-CCP-treated individuals were generalizable and could predict poor clinical outcomes for all trial participants regardless of CCP status, we used the above PLS-R model that used only the LASSO-selected S-specific features (Figure I) to predict whether participants regardless of CCP treatment could be classified into 1) high severity COVID-19 outcome (CSC > 20) or 2) low severity COVID-19 (CSC <= 20). The model was highly predictive of disease severity achieving an average AUC in an ROC curve of 77% (Figure 2K) and demonstrated that these six inflammatory S antibody features predicted worse COVID-19 clinical outcomes. This reinforces that the inflammatory S antibody features in control participants are associated with more severe outcomes. Thus, here we show that CCP modulation of S humoral immunity and dampening of inflammation are linked to improved disease outcomes.

### Correction for co-morbidities points to a robust Nucleocapsid-specific antibody signature of CCP-treatment

A major challenge in understanding the effect of CCP relates to the heterogeneity of COVID-19 clinical disease. Co-morbid conditions including obesity, diabetes, cardiovascular disease, chronic kidney disease, concomitant immunosuppression, and cancer have all been associated with more severe COVID-19.(Prevention, 2021) Age and obesity have both been associated with decreased B cell responses and lower antibody responses to pathogens and vaccines.(Frasca et al., 2017) Thus, to account for these covariates in our analysis of CCP-induced humoral immune evolution, we used a nested mixed-linear modeling approach of the antibody profiles of CCP-treated and control participants over the first 15 days of the study (Day 1 to Day 15 of the Clinical Trial). Age, sex, race, ethnicity (Latinx vs. non-latinx), blood type, quarter of enrollment, diabetes, cardiovascular disease, hypertension, obesity, chronic kidney disease, cancer, prior immunosuppression, concomitant treatment with remdesivir at study entry, concomitant treatment with steroids at study entry, and time of symptom onset were included in the models. For each antibody feature, we generated two mixed linear models, one with treatment group (CCP-treatment vs. control) incorporated as a fixed effect (the full model) and the other model without the treatment group (null model). Then, we compared the two nested models with the likelihood ratio test (LRT) to quantify whether treatment group information improved the model fitting, thus identifying Ab features whose trajectories are affected by CCP treatment. We then extracted the T values (normalized coefficient) of treatment group variable in the full mixed linear model to quantify the magnitude of effect CCP treatment has on Ab features. Altogether, antibody features significantly affected by CCP-treatment were defined as having a T value > 2 and a two-sided p-value < 0.05. Most antibody features that significantly differed between CCP-treated and control individuals were enriched in CCP-treated individuals (Figure 3A), suggesting that many of the S-specific features increased in control individuals in Figure 2 F are influenced by known COVID-19 disease severity risk factors. Intriguingly, most features enriched in CCP-treated individuals were N-specific antibody features including binding strength to N-specific-FcγR2B, -FcγR3B, and -ADCD. Further, the only feature enriched in control participants (T-value < −2) was RBD antibody binding to the IgA Fc-receptor FcαR (Figure 3A), also identified in our modeling based on days from symptom onset (Figure 2F). To ensure the validity of the model results, we next confirmed that the N-feature levels prior to randomization were balanced across the two arms (Figure 3B, Supplementary Figure 3A). Thus, using a multivariate mixed-effects model, a robust and unexpected N-specific humoral signature emerged in CCP-treated participants.

**Figure 3:**
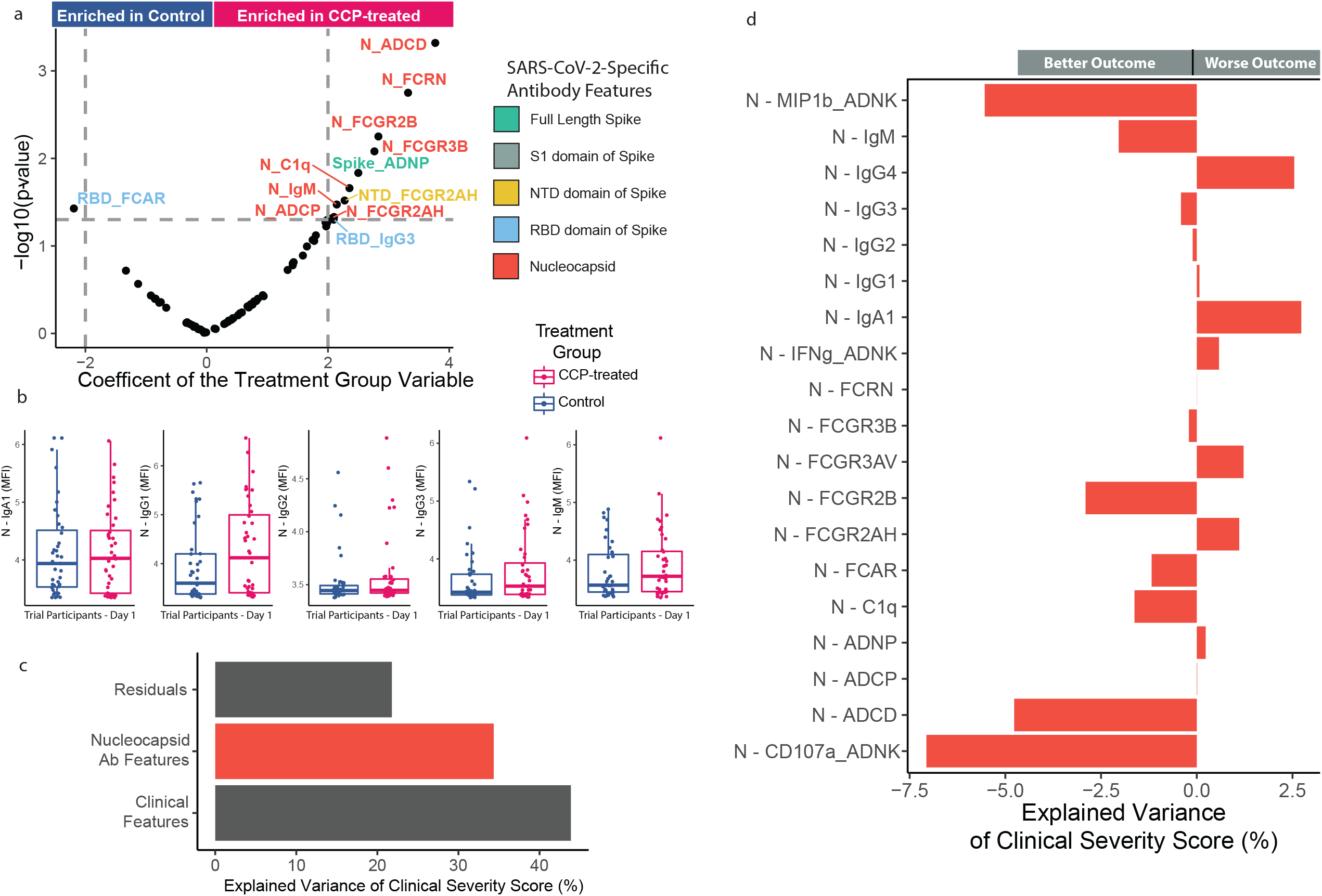
CCP also provides clinical benefit by enhancing nucleocapsid-focused humoral response. A nested mixed linear model was created for each antibody feature with and without a variable accounting for patient’s treatment group (CCP treatment vs. control) to assess CCP’s effect on the host anti-SARS-CoV-2 humoral development. (a) Volcano plot shows the T-value (normalized coefficient) of the patient treatment group variable incorporated in the mixed linear model (x-axis) and p-value of the likelihood ratio test for the model fit difference between the two nested models (y-axis). A positive T-value represents a feature enriched in CCP-treated individuals and a negative T-value represents a feature enriched in control individuals. (b-c) Nucleocapsid-specific humoral profile of CCP-treated and control patients (b) Box plots of selected Nucleocapsid-specific features prior to treatment (Day 1). Each box represents the median (central line) and IQR (25% and 75% percentiles) and the two whiskers represent 1.5 * IQR. (c-d) A Linear regression model was used to assess if nucleocapsid features could predict COVID-19 clinical severity of CCP-treated and control patients as measured by the clinical severity score. (c) The bar plot shows the percentage of explained variance (%) by clinical data (clinical characteristics, severity risk factors, and concurrent medications) and Nucleocapsid antibody features. (d) The bar plot shows the contribution of each Nucleocapsid antibody feature to COVID-19 clinical severity. The magnitude represents the percentage of variation in the clinical severity score explained by each feature, and the directions represents whether the antibody feature was associated with better (i.e. negative Explained Variance of Clinical Severity Score (%)) or worse (i.e. positive Explained Variance of Clinical Severity Score (%)) clinical outcomes.

### Nucleocapsid features are associated with improved outcomes in CCP-treated and Control participants

Given the enrichment of N-specific antibody features in CCP recipients, we next sought to understand the relationship between N-specific antibodies and clinical outcomes in all study participants. To define whether certain N-specific antibody features were associated with clinical outcomes based on CSC, we applied a linear effects model to the N-specific antibody profiles of CCP-recipients and controls over the first two weeks of the trial (Day 1 to Day 15). Data were corrected for co-morbidities associated with COVID-19 disease. N-specific features explained 30% of the variation in clinical outcome across the cohort (Figure 3C). Additionally, the association of individual N-specific antibody features and clinical outcome (CSC score) (Figure 3D) pointed to an association between most N-specific antibody features and better clinical outcomes. Specifically, N-specific ADCD, the most strongly CCP-enriched antibody feature (Figure 3A), was also one of the most strongly associated with better clinical outcome (Figure 3D). N-specific FcγR2B, FcγR3B, C1q binding, and IgM titers were all also enriched in CCP-treated individuals and associated with improved clinical outcomes (Figure 3A, 3D). On the other hand, N-specific NK CD107a and MIP1b expression were most strongly associated with better outcomes (Figure 3D), though not differentially enriched between CCP-treated and control participants (Figure 3A, 2F), suggesting that N-specific ADCC may be beneficial in COVID-19 but not affected by CCP treatment. Further, not all N-specific antibody responses are beneficial. N-specific IgA and IgG4 levels were not enriched in CCP-treated individuals and were associated with worse outcomes. Thus, these analyses showed that CCP treatment-associated N-specific humoral immune responses are associated with clinical benefit.

### COVID-19 participants with low functional antibodies benefited most from CCP treatment

Emerging data from monoclonal therapeutics trials suggest that participants who have not yet generated an antibody response to SARS-CoV2 may benefit the most from monoclonal antibody therapy.(Group et al.; Herman et al., 2021a; Libster et al., 2021; Weinreich et al., 2020) Next, to understand which participants benefited the most from CCP therapy in this study, participants were clustered based on their Day 1 SARS-CoV-2 antibody profiles using a Spearman correlation distance-based Neighborhood clustering approach (Figure 4A). Four clusters of participants with similar pre-existing antibody profiles appeared (Supplementary Figures 4A, B). Cluster 1 contained participants with the highest S and N-specific humoral responses, while Clusters 2, 3, and 4 had more varied antibody profiles. Cluster 4 included individuals with the lowest S and N titers across all antibody features (Figure 4A). Principal component analysis and co-correlation network structure demonstrated that Cluster 1 and 4 were most distinct in their SARS-CoV-2 antibody profiles (Figure 4B, Supplementary Figure 4B). Strikingly, CCP-treated participants in Cluster 4 exhibited the greatest benefit (lower CSC) compared to control participants (Figure 4C,F). To gain a granular sense of how Cluster 4 individuals differed from the other clusters, we performed univariate testing comparing Cluster 4 antibodies profiles to the antibody profiles of non-Cluster 4 participants, including Cluster 1,2,3, (Figure 4D, E). Specifically, Cluster 4 participants possessed lower S- and N-specific antibody functions, most strikingly exhibiting the lowest S and N-specific ADCP, low S- and N-specific Ab-mediated NK cell MIP1b production as well as lower S- and N-specific IgA1 and IgG1 titers (Figure 4D, E, Supplementary Figure 6). Thus, these results suggested that participants with lower functional antibody responses were more likely to benefit from CCP treatment.

**Figure 4:**
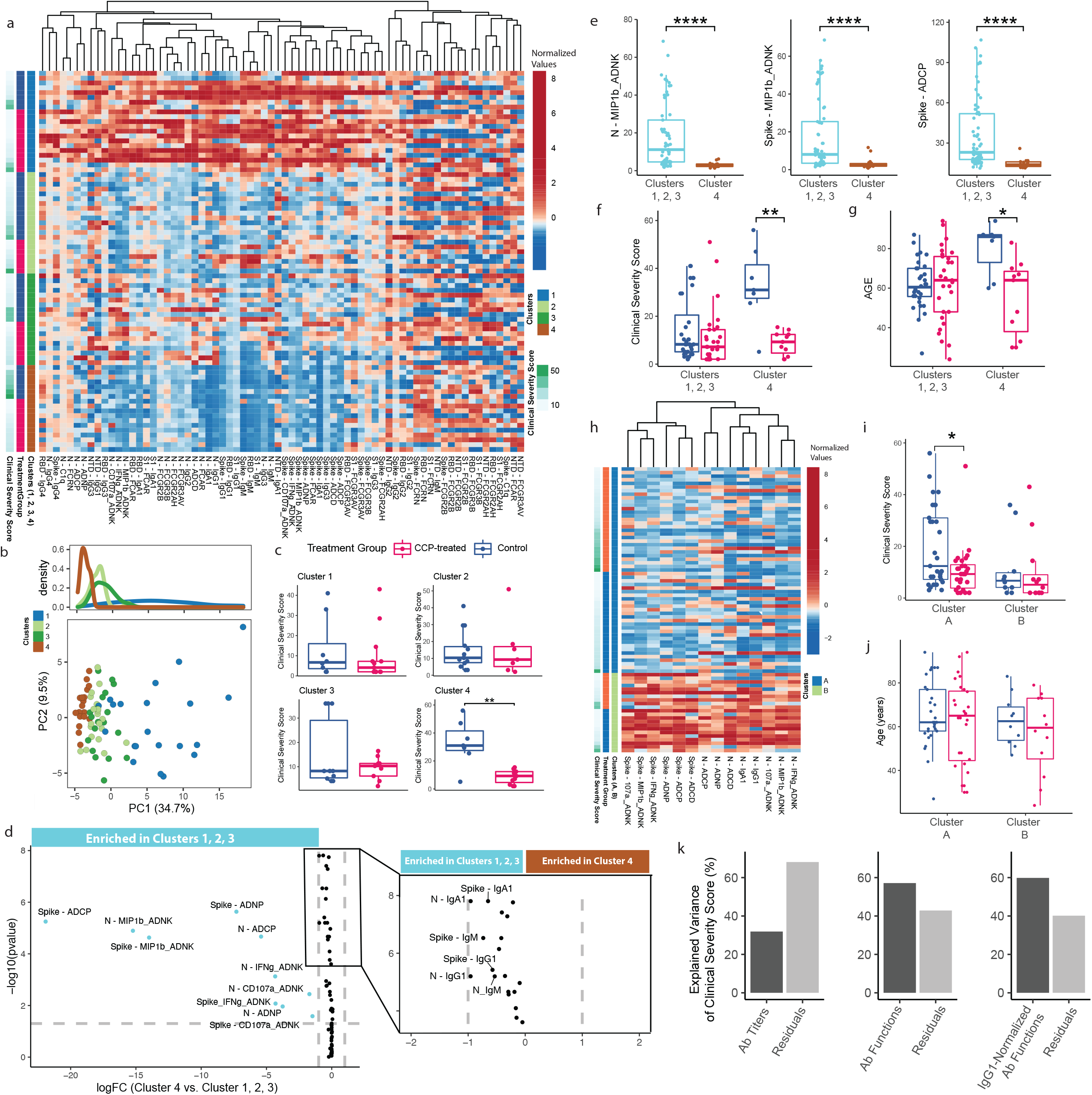
Patients with less functional pre-existing antibodies benefit the most from CCP. Seventy-nine samples collected before treatment (Day 1) were used to evaluate the association between antibody profiles and clinical severity, as measured by the clinical severity score at day 28. Spearman correlation-based clustering to identify population benefiting from CCP (A-F). (A) The heatmap represents the normalized Day 1 antibody profiles. Patient samples were clustered into four groups based on the similarity of spearman correlation coefficients of Day 1 SARS-CoV-2 antibody profiles between samples. (B) PCA plot (bottom) shows the relatedness of patients in each of the identified four clusters and the density plot (top) displays the organization of the patient samples from each of the four clusters along principal component 1 (PC1). (C) The box plots show the clinical severity scores of CCP-treated and control patients in each of the four clusters. A Wilcox Rank test was used to test for differences in clinical severity scores between the two groups in each cluster (two-sided p value: 0.365, 0.799, 1, 0.00415). (D) The volcano plots show the antibody function, titers, and Fc receptor binding features that were most different between Cluster 4 and the rest of the population (Cluster 1, 2, 3). The pop-out highlights features with a log fold-change (logFC) between −2 and 2 as well as a p-value < 0.05. The x-axis represents the log-fold change of Cluster 4 over Cluster 1,2,3 and the y-axis represents the p-value from a two-sided Wilcoxon test. Each dot represents an antibody feature. Ab features with |logFC|> 2 were colored by the population in which they were enriched, i.e. in Cluster 4 (brown) or in Cluster 1, 2,3 (cyan). (E) Box plots of representative antibody functions enriched in non-cluster 4 patients. (F) Boxplot of the Clinical Severity Score of CCP-treated and control patients in Cluster 4 and Cluster 1, 2, 3. A two-sided Wilcoxon test was performed to compare age between treatment arms. (G) Boxplot of the age of CCP-treated and control patient in Cluster 4 and Cluster 1, 2, 3. A two-sided Wilcoxon test was performed to compare age between treatment arms. (H-J) Re-clustering patients based on benefit signature identified in (D, E) on the whole population. (H) The heatmap shows the two clusters (Cluster A and B) identified by the benefit signature – the features that most distinguished Cluster 4 from Clusters 1, 2, 3 patients (the top 12 features). (I) Box plots of clinical severity of CCP-treated and control groups in cluster A and B. The difference between CCP-treated and control patient’s clinical severity was tested by the two-sided Wilcoxon test. (J) Box plots of age of CCP-treated and control groups in cluster A and B. (K) Three separate linear regression models were used to assess which type of pre-existing antibody features best predicted clinical severity in CCP-treated individuals. The bar plots show the percentage of explained variance (%) by antibody titers, antibody functions, or IgG1 titer-corrected antibody functions in the separate models. We used the top 12 features that differed between Cluster 4 and Cluster 1, 2, 3 for the linear regression model of each antibody feature category. For the boxplots in C, E, F, G, I, and J each box represents the median (central line) and IQR (25% and 75% percentiles) and the two whiskers represent 1.5 * IQR. * represents a p-value < 0.05. ** represents a p-value <0.01. *** represents a p-value <0.001. **** represents a p-value < 0.0001.

Additional comparisons of clinical factors across the 4 clusters pointed to relatively balanced symptom duration prior to trial enrollment (Supplementary Figure 4C). However, participants in Cluster 4 were less likely to have chronic kidney disease (CKD) or be obese, were less likely to be African American, and were more likely to have enrolled later in the clinical trial period (May 2020 through January 2021) (Supplementary Table 1). Further, Cluster 4 controls were significantly older than the Cluster 4 CCP-treated group (Figure 4G, Supplementary Table 2). To further define whether the CCP-response signatures identified at baseline in Cluster 4 could predict benefit from CCP across the whole trial, we re-clustered participants based on the CCP benefit signature, using all N- and S-specific antibody functional measurements and all antibody titers with |log FC| > 0.75 (N-IgG1 and N-IgA1). Antibody profiles clustered into 2 groups (Figure 4H, Supplementary Figure 4D, E): Cluster A consisted of a heterogenous mix of participants with overall lower S- and N-antibody features (Figure 4H) and with statistically significant lower CSC (and better clinical outcome) in CCP-treated participants (Figure 4F). In contrast, Cluster B consisted predominately of higher levels of S-and N-specific antibody functions and titers (Figure 4H) and nearly identical CSC across CCP-treated and control participants (Figure 4I). Because clinical characteristics were equally distributed across the two-cluster model (Supplementary Table 3) as well as across Cluster A CCP-treated and control groups (Supplementary Table 4), these data suggest that the quality of the pre-existing humoral immune response to SARS-CoV-2 infection largely explained the benefit they received from CCP rather than patient demographic factors or COVID-19 severity risk factors.

We next sought to identify specific pre-existing antibody functions or levels that were predictive of benefit from CCP treatment. We created three linear models that predicted the clinical severity measured by the CSC of CCP-treated participants based on either their pre-existing antibody levels, unadjusted antibody functions, or IgG1-normalized antibody functions. For this comparison, we used the top 12 antibody functions and the top 12 antibody levels that differed between Cluster 4 and Cluster 1,2,3 (Figure 4D). Antibody levels alone only predicted 32% of the variation in CSC, while antibody functions predicted 57% percent of variation in the CSC (Figure 4K). Further when we normalized the antibody functions by IgG1 to eliminate the influence of antibody level differences, we continued to explain 60% of the variation in CSC (Figure 4K). On an individual feature level, S titers contributed more to the explained variance but S and N-specific humoral features both contributed to the explained variance of the functions (Supplementary Figures 4F, G, H). Thus, we found that pre-existing anti-SARS-CoV-2 humoral functions are a stronger predictor of response to therapy than seronegative status alone.

### Two months later, CCP-treatment resulted in a sustained shift in the inflammatory status of Spike-specific antibodies via glycosylation changes

Based on the pharmacokinetics of IVIG in secondary immunodeficiencies, it is unlikely that antibodies from 2 units of CCP (∼400mL) would continue to circulate for more than a month after therapy.(Koleba and Ensom, 2006) Thus, we next examined whether CCP exerted long-lasting effects on recipient’s SARS-CoV-2 humoral immune response. First, we found that CCP-treated and control participants did not have significantly different Spike IgG1 levels (Figure 5A). In addition to changes in the overall levels of antibodies, the functional and inflammatory properties of antibodies are regulated by changes in IgG Fc-glycosylation at Asparagine 297.(Arnold et al., 2007; Jennewein and Alter, 2017; Raju, 2008) Given the importance of the Fc-glycan in severe COVID-19(Chakraborty et al., 2020; Larsen et al., 2020), we profiled Fc glycan differences across CCP-treated and control participants two months after treatment (Day 60). CCP-treated individuals exhibited a selective enrichment of Spike-specific disialylated and diglycosylated peaks such as G2S2F, G2S2B, G2S21F (Figure 5 B, C). A LASSO/PLS-DA model, using S-specific Fc-glycans profile features only, was able to separate CCP-treated from control participants (Figure 5D and Supplementary Figure 6A). Among the Fc-glycan structures, G2S2F and G2S1B were selectively enriched in CCP-treated, while G1FB.G2 was enriched in control participants (Figure 5E). Moreover, a co-correlational network was constructed to gain deeper insights into the collection of Fc-glycans that may co-evolve in the setting of CCP treatment (Figure 5F). G2S2F, enriched in CCP-treated participants, was strongly correlated with Sialyation, Disialyation, digalactosylation as well as individual digalactosylated Fc glycan species including G2S1FB, G2S2, and G2S2FB. Conversely, G2S2F was strongly anti-correlated with monogalactosylation and asiaylated features such as G2F, G1F.1FB, and G1FB.G2 pointing to an enrichment of heavily sialylated and galactosylated S antibodies in CCP-treated individuals. As both high sialyation(Anthony et al., 2008; Kaneko et al., 2006) and galactosylation (Karsten et al., 2012) have been linked to anti-inflammatory antibody activity, these data point to the evolution of anti-inflammatory Spike-specific antibody profiles following CCP therapy. A second network was observed including the CCP-treated enriched feature G2S1B with another bisected feature G2B, point to a potential role for bisecting GlcNAc in CCP-treated individuals. Both G2S2FB and disiaylated antibodies were individually significantly enriched in CCP-treated individuals compared to non-treated controls (Figure 5 G, H). Together, these data point to a longer-term effect of CCP on shaping the inflammatory profile of the evolving SARS-CoV-2 humoral immune response.

**Figure 5:**
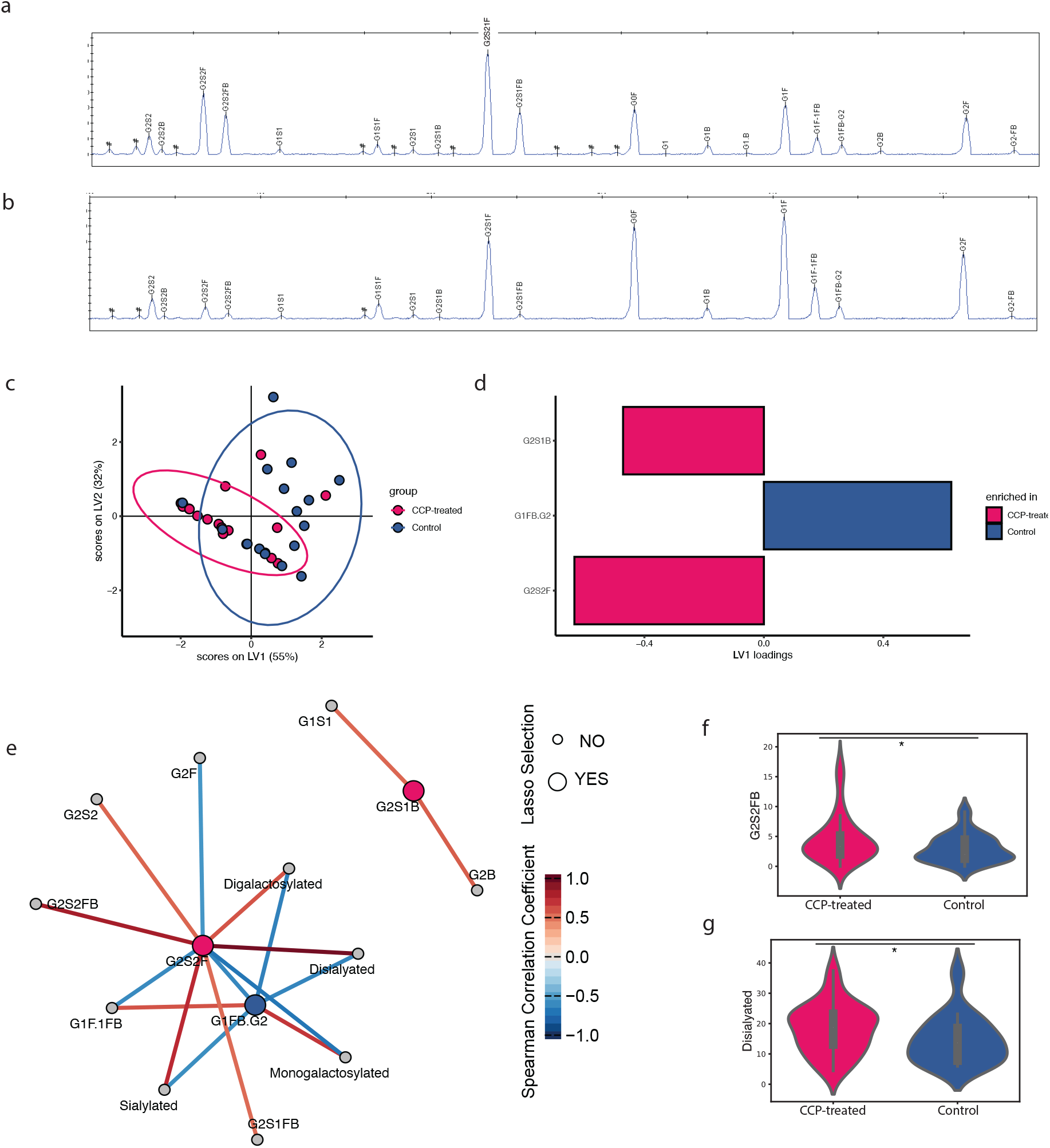
CCP Recipients have highly sialylated and galactosylated S-specific Fc modifications long after treatment. (A) Bar graphs of Spike-specific IgG1 levels in CCP-treated and control participants 60 days after randomization. S-Specific Fc glycosylation patterns were measured by capillary electrophoresis in all participants with D60 samples collected from CCP-treated (n=19) and Control (n=16) participants. Representative chromatographs of CCP-treated (B) and Control (C) participants. (D, E) LASSO PLS-DA was performed to identify the Fc glycan features that separated the two groups. The PLS-DA scores plot (D) shows that the S-specific Fc-glycans can separate CCP-treated from control participants with LV1 explaining 41% of variation that separates the two groups along the x-axis. Each dot shows an Fc glycan measurement. (E) LV1 loadings plot shows the LASSO-selected features. Pink represents features enriched in CCP-treated participants and Blue represents features enriched in Control participants. (F) The Spearman correlation network shows the co-correlated features (small nodes) that are significantly correlated (p-value < 0.05 after multiple test correction using the Benjamini-Hochberg procedure, spearman correlation coefficient > 0.5) with the model selected features (large nodes). Large nodes are colored by the treatment arm in which they are enriched. Edges are colored by magnitude and sign of correlation, with dark red and dark blue representing strong correlation and anti-correlation respectively. (G,H) Univariate plots for G2S2FB (G) and Disialylated (H) Fc glycans in CCP-treated (Pink) and Control (Blue) participants. * represents a p-value < 0.05 by a two-sided Wilcoxon rank test.

### Nucleocapsid immunodominance persists two months after CCP-treatment

Given the presence of a persistent anti-inflammatory humoral signature on S-specific antibodies two months after treatment, we finally aimed to define whether early signatures of response to therapy persisted over time. Thus, we investigated the SARS-CoV-2 specific antibody profiles of CCP-treated and control participants two months after treatment. Two months after therapy, CCP-treated individuals continued to exhibit enhanced N-specific antibody titers and Fc-receptor binding antibodies, while control participants still had higher S1- and RBD-specific antibody titers and Fc-receptor binding (Figure 6 A, B), pointing to the persistence of the immunodominant shift associated with CCP therapy. Furthermore, using a LASSO/PLS-DA, CCP-treated individuals continued to exhibit a unique overall humoral immune profile compared to control participants (Figure 6C-E). Only 4 of the total 70 features, collected per plasma sample, were sufficient to separate out antibody profiles across the 2 groups, 2 months after therapy. The 4 features included NTD-specific IgA1 and NTD-specific FcγR3A binding antibody levels that were enriched in control participants in our model (Figure 6D) and N-specific IgM and C1q binding antibodies were selectively enriched in CCP-treated individuals (Figure 6D). Moreover, the LASSO-selected feature co-correlation network highlighted the presence of additional S-specific features associated with NTD-specific FcγR3AV binding levels in controls. Interestingly, these features were inversely correlated with N-specific ADCP, further highlighting the dichotomous response represented by either a S- or an N-focused antibody profiles (Figure 6F). Further, the CCP-treatment enriched feature N-C1q was tightly co-correlated with 15 other N-specific antibody features that were selectively enriched among the CCP treated individuals. The tight correlation of N-specific ADCD with and C1q (Figure 6F) as well as the LASSO selection of N-specific C1q and N-specific IgM (Figure 6D) suggested that CCP treatment may contribute to a durable classical complement pathway-focused response to N two months after CCP treatment. These data point to durable effects of CCP that result in long-lived attenuation of S-specific inflammatory responses in favor of a durable N-specific complement-focused humoral response. Collectively, these data suggest that the benefit of CCP in hospitalized COVID-19 patients is due in part to an immunodominance shift in humoral immune evolution, marked by a reduced S-specific humoral immune responses and augmented N-specific humoral immunity, resulting in durable changes in antibody profiles months after treatment.

**Figure 6:**
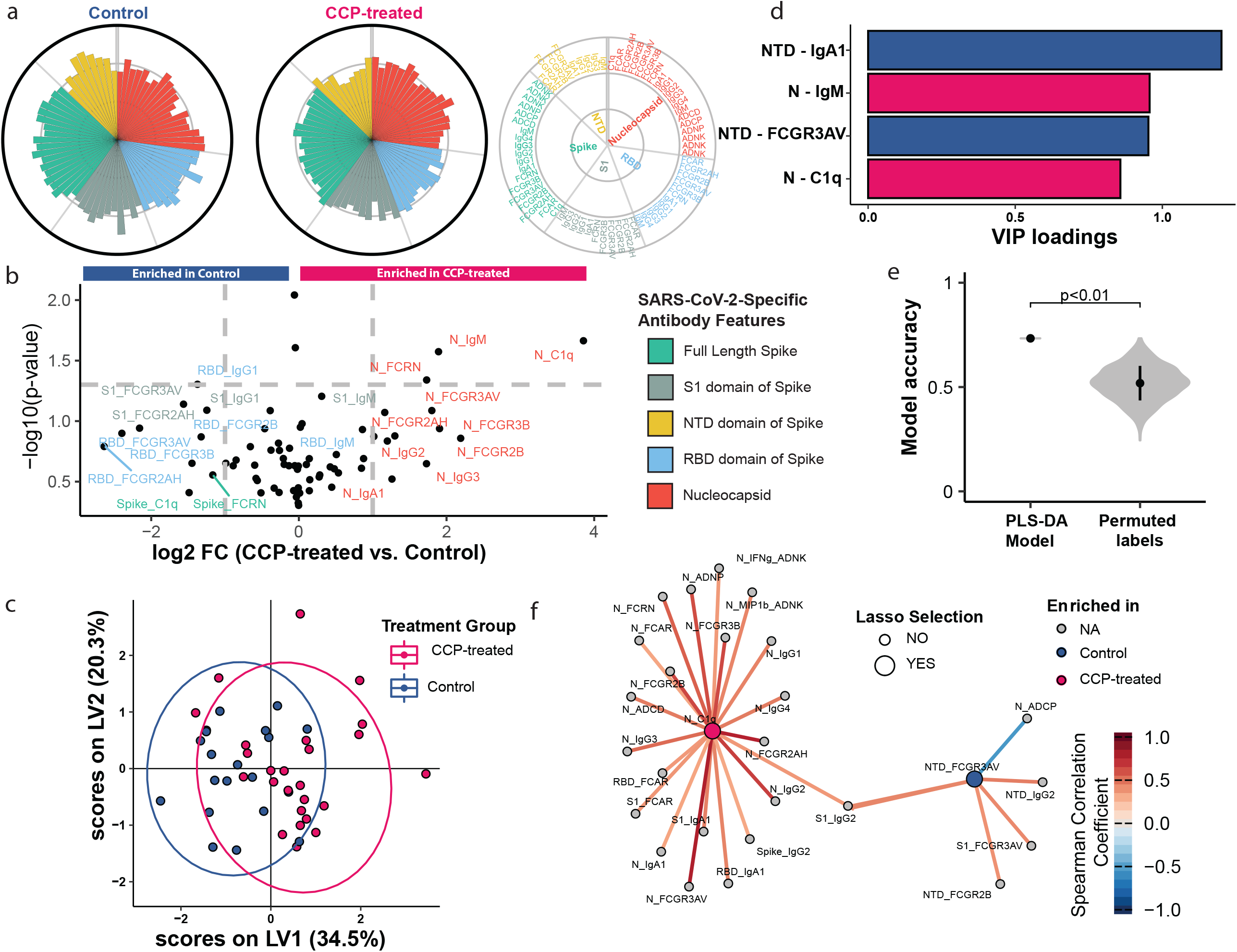
Long-lasting Nucleocapsid immunodominance in CCP on recipients. SARS-CoV-2 functional antibody profiles of forty-five patients (CCP-treated n=25, control n=20) on Day 60. (A) Polar plots of the mean percentile of each antibody feature across the control (left) and CCP-treated study arms (right). The features were grouped by the antigen detectors and are depicted in a key. (B) Volcano plot showing the difference between the humoral profile of CCP-treated group and control group by the fold change of mean value (x-axis) and two-sided p-value from Wilcoxon rank test (y-axis). (C-F) LASSO PLS-DA model identified the antibody features that distinguish CCP-treated from control patients at day 60. (C) The PLS-DA score plot demonstrates that CCP-treated and control Day 60 patients can be discriminated by the LASSO selected features. Each dot shows an individual patient. (D) Variable importance in projection (VIP) score of the selected features. The magnitude indicates the importance of the features in driving separation in the model. Pink represents a feature enriched in CCP-treated and blue represents a feature enriched in control patients. (E) The performance and robustness of the model was validated with permutation testing. The violin plot shows the distributions of repeated classification accuracy testing using label permutation. The p-value from the permutation testing is two-sided. Black squares indicate the median accuracy and black lines representing one standard deviation. (F) The correlation network shows the co-correlated features (small nodes) that are significantly correlated (p-value < 0.05 after multiple test correction using the Benjamini-Hochberg procedure, spearman correlation coefficient > 0.3) with the model selected features (large nodes). Large nodes are colored by the treatment arm in which they are enriched.

## Discussion

Since the start of the COVID-19 pandemic, many clinical trials have studied the efficacy of CCP. Many large studies of hospitalized COVID-19 patient have not demonstrated efficacy from CCP, however a select number of studies of high-titer CCP earlier in disease have shown benefit.(Avendaño-Solá et al., 2021; Bar et al., 2021; Group et al.; Investigators et al., 2021; Joyner et al., 2021; Libster et al., 2021; O’Donnell et al., 2021; Sullivan et al., 2021; Yoon et al., 2021) Here we applied systems serology to a randomized study showing clinical benefit of CCP treatment early in hospitalization for COVID-19 pneumonia(Bar et al., 2021) to understand the signatures of protective immunity provided by CCP that could inform future antibody therapeutic development. We found that CCP shifted immunodominance to SARS-CoV-2 by diminishing the S-focused evolution in exchange for expanded N-specific activity. The clinical benefit associated with this immunodominance shift suggests three major findings: 1) the importance of blunting the inflammatory S-targeted humoral response in severe COVID-19, 2) the critical role of clearing N-specific immune complexes, and 3) the anti-inflammatory effects on the S and N humoral response are long lasting. These findings further extend our previous study examining a cohort of patients treated with CCP early in the COVID-19 pandemic where CCP enriched in N-specific antibodies blunted the development of inflammatory anti-SARS-CoV-2 host response, but was not designed to assess the clinical benefit of CCP.(Herman et al., 2021b) Here in this study, we found that CCP treatment led to lower levels and slower development of FcR2AH, the predominant FcR on monocytes, and binding of S-, S1-, and RBD-specific antibodies, suggesting CCP may blunt the development of monocyte-activating S antibodies. Emerging work has shown that the afucosylated inflammatory Spike antibodies found in participants with severe COVID-19 promote macrophages to produce pro-inflammatory cytokines(Hoepel et al., 2021) and inflammatory monocyte/macrophages that are central to the hyperinflammatory state in severe COVID-19.(Zhou et al., 2020) This strongly argues that part of the therapeutic benefit of a polyclonal antibody therapy is its capacity for immunomodulation. For CCP treatment of hospitalized patients with COVID-19, this effect manifests as a dampening of the antibody-induced macrophage/monocyte hyperinflammatory host response acutely and possibly even months after the resolution of COVID-19 viremia.

Strikingly, our results show that an N-focused immunodominance in COVID-19 disease is associated with better clinical outcomes. Emerging work suggests freely circulating N protein can activate complement via the alternative pathway(Gao et al., 2020; Kang et al., 2021) and is likely involved in the hyperinflammatory lung damage through which severe COVID-19 leads to acute respiratory distress syndrome.(Gao et al., 2020; Ma et al., 2021) Further, monoclonal antibodies targeting N in an in vitro system can inhibits free N-induced MASP-2 activation.(Kang et al., 2021) In this work we found that CCP induces stronger N-specific humoral responses, which were associated with improved clinical outcomes. This suggests that N immunodominance may be a mechanism to attenuate inflammatory activity of N-specific immune complexes in the lung, while allowing the rest of the immune system to control and clear the infection. Further, N Ab binding is not therapeutic on its own. Of the nineteen N-specific antibody features measured in this work that were associated with CCP treatment, the two most strongly associated with clinical benefit were Antibody-dependent complement deposition and FcR2B binding. These functional correlates provide hints on how to make a new generation of anti-N therapeutics to dampen inflammation in severe COVID-19. Importantly, we also found that months later, CCP-treated individuals had N-dominance humoral profiles. Hence, the shift to N immunodominance was long lasting. Together, the long-lasting immunodominance shift associated with CCP treatment identified in this work highlights the potential importance of clearing N-immune complexes early in severe COVID-19.

The observed immunodominance shifts associated with CCP treatment were linked to improved clinical outcomes. Using orthogonal analytic approaches, we found that the diminished S-features and enhanced N-features in CCP-treated individuals were associated with better outcomes in control as well as in CCP-treated participants. Thus, our data suggest that the benefit of CCP in hospitalized COVID-19 patients may not only be due to neutralizing antibody but also through shifting immunodominance of CCP recipients’ immune response via antibody functional activity. This novel immunomodulatory possibility suggests that passive antibody therapy may have distinct benefits in COVID-19 patients as compared to anti-RBD monoclonal antibodies and antivirals such nirmatrelvir/ritonavir(Pfizer, 2021) and molnupiravir,(Bernal et al., 2021) both of which target viral invasion/replication to mediate a clinical benefit. Furthermore, the continued evolution of new variants of concern (VOC), most recently omicron, has led to a loss of activity for many of the RBD-targeted monoclonal antibodies.(Aggarwal et al., 2021; Cameroni et al., 2021; Cao et al., 2021; Planas et al., 2021) Polyclonal antibody therapies such as CCP, hybrid convalescent/vaccinated plasma(Focosi et al., 2021), COVID-19 hyperimmunoglobulin,(Ali et al., 2021) equine COIVD-19 hyperimmunoglobulin, and transchromic COVID-19 hyperimmunoglobulin such as SAB-185(Liu et al., 2021) may contain the breadth of antibodies needed to combat future VOCs by targeting multiple epitopes on the RBD, multiple sites within Spike, and multiple proteins(Tang et al., 2021) within the SARS-CoV-2 proteome.

Who among hospitalized patients should receive passive antibody therapy remains an unsettled question in COVID-19 clinical care. Many lines of evidence suggest that anti-S monoclonal antibodies(Group et al., 2021) and polyclonal high-titer CCP(Libster et al., 2021) are more effective when given earlier in COVID-19 disease, especially to seronegative patients. Here we found that both S and N antibody functions were better predictors of clinical benefit from CCP than antibody titer or duration of symptomatic COVID-19. Strikingly our CCP benefit signature, chosen by an un-biased clustering approach, did not include any S or S sub-domain features and was still able to identify the CCP-participants who had better outcomes than pre-existing antibody-matched controls. These findings suggest that for hospitalized patients with moderate to severe disease, antibody quality and function were more important than antibody quantity to mediate a clinical benefit. Further, selecting recipients of passive antibody therapy strictly by S antibody level, we may miss patients with high levels of low-quality antibodies who could benefit from CCP or monoclonal antibody therapy.

In this work, we studied a randomized control clinical trial of hospitalized COVID-19 patients with severe disease where CCP treatment led to a significant decrease in mortality and improvement in a COVID19 disease severity. Though the UPENN trial enrolled fewer participants than the multicenter CCP trials, it used local, single sourced plasma, which may in part explain its positive results.(Kunze et al., 2021) By focusing our analysis on this single center, we were able to use time-to-event analysis as part of the CSC and help us better parse out the continuum of COVID-19 outcomes. Designed as a randomized trial that compared CCP to standard of care, our analysis of the UPenn CCP2 trial cannot rule out the effects of non-antibody proteins present in convalescent plasma. However, Sullivan et al. found CCP reduced risk of hospitalization in a trial of CCP vs. fresh-frozen plasma (FFP), suggesting that serum proteins present in both FFP and CCP are not responsible for the clinical benefit found in their and our trial.(Sullivan et al., 2021) Further, we were not able to seek CCP-specific antibody features that drove COVID-19 clinical outcomes since the majority of participants in our clinical trial received CCP from two separate donors. Additionally, this study was conducted at a time before widespread vaccination in the United States, thus we do not know how vaccination status will affect CCP response. Since the majority of participants in this study were already being treated with corticosteroids and remdesivir, we cannot address whether the activity of CCP we found here is independent or contingent on combination treatment. Despite these limitations, we were able to use deep humoral immune profiling to understand how CCP modulates host immunodominance and provides clinical benefit. Expanding this approach in larger clinical trials will be essential to validate these findings.

The emergence of the omicron variant has rendered most of our monoclonal antibody therapeutics no longer active.(Aggarwal et al., 2021; Cameroni et al., 2021; Cao et al., 2021; Planas et al., 2021) Now there is a renewed interest in use of polyclonal antibody therapies like CCP, which are less likely to loose efficacy to new variants because they target multiple sites in the virus and plasma from survivors of recently circulating variants can be procured relatively quickly. By using a systemic approach to study correlates of therapeutic benefit of CCP, we have found novel targets for future severe COVID-19 disease modifying treatments. Our findings contribute to a burgeoning literature showing the promise of anti-N monoclonal antibodies as disease modifying treatment for severe COVID-19 induced hyperinflammation. Further, our findings show that by choosing CCP based on high S titers alone and selecting patients based on low pre-existing S titers, we are likely incorrectly matching patients with therapies. Finally, our research confirms the importance of the functional S and N antibody response in treatment of COVID-19 disease and should guide development of COVID-19 monoclonal and polyclonal antibody therapeutics that focus not only on neutralization, but also on Fc-directed functionality.

## Methods

### Clinical Cohort

The cohort described here participated in a randomized control trial of convalescent plasma in hospitalized patients with severe COVID-19, as described in Bar et al.(Bar et al., 2021) Briefly, the study enrolled hospitalized adults with RT-PCR-confirmed SARS-CoV-2 infection, radiographic documentation of pneumonia, and abnormal respiratory status, defined as room air saturation of oxygen (SaO2) <93%, or requiring supplemental oxygen, or tachypnea with a respiratory rate ≥30 breaths per minute. Participants were excluded if they had a contraindication to transfusion, were participating in other clinical trials of investigational COVID-19 therapy, if there was clinical suspicion that the etiology of acute illness was primarily due to a condition other than COVID-19, or if ABO-compatible CCP was unavailable. Between May 2020 and January 2021, a total of 80 eligible participants were randomized to receive either 2 units of CCP and standard of care (treatment arm) versus standard of care alone (control arm). Participants were assigned to treatment or control in a 1:1 ratio. 41 participants were randomized to treatment, but two declined CCP administration and 40 were included in our analysis; 39 participants were randomized to control. 39 participants in the treatment arm received up to 2 units of convalescent plasma on study day 1; with 4 participants receiving 2 units from the same donor and 35 receiving units from two distinct donors. Participants were enrolled a median of 6 days (IQR 4 – 9) after the onset of COVID-19 symptoms. None of the participants were on mechanical ventilation on enrollment. The majority of participants received steroids (83%) and remdesivir (81%) at enrollment. The median age of participants was 63 (IQR 52 – 74), and 41% had diabetes, 67% had hypertension, 45% had obesity, 32% had chronic kidney disease, 27% had cancer, and 14% had immunodeficiencies. Of the enrolled participants, 54% were female and 45% were male. The majority of participants identified as African American (53%), with 5% identifying as Asian, 4% Identifying as Latino/a, 34% identifying as Non-Latino/a Caucasian, and 4% without an identified race or ethnicity.

### Antibody Titer and Fc-Receptor Binding Assays

Antigen-specific antibody subclass, isotype, and Fc-receptor (FcR) binding levels were assayed with a customized multiplexed Luminex bead array, as previously described.(Brown et al., 2017) This allowed for relative quantification of antigen-specific humoral responses in a high-throughput manner and simultaneous detection of many antigens. A panel of SARS-CoV-2 antigens including the full spike glycoprotein (S) (provided by Lake Pharma), receptor binding domain (RBD) (Provided by Aaron Schmidt, Ragon Institute) nucleocapsid (N) (Aalto Bio Reagents, Dublin, Ireland), S1 (Sino Biological, Beijing, China) S2 (Sino Biological, Beijing, China), and N-terminal domain (NTD) (Sino Biological, Beijing, China) were used. Control antigens were run including a mix of three Flu-HA proteins (H1N1/A/New Caledonia/20/99, H1N1/A/Solomon Islands/3/2006, H3N2)(A/Brisbane/10/2007 – Immune Tech) and Ebola glycoprotein (IBT Bioservices). In brief, antigens were coupled to uniquely fluorescent magnetic carboxyl-modified microspheres (Luminex Corporation, Austin, TX) using 1-Ethyl-3- (3-dimethylaminopropyl) carbodiimide (EDC) (Thermo Fisher Scientific, Waltham, MA) and Sulfo-N-hydroxysuccinimide (NHS) (Thermo Fisher Scientific, Waltham, MA). Antigen-coupled microspheres were then blocked, washed, and incubated for 16 hours at 4°C while rocking at 700 rpm with diluted plasma samples at plate concentrations of 1:12,000 for all subclasses and isotypes and C1q and FcRn binding and 1:120,000 for all other Fc-receptors to form immune complexes in a 20 uL volume in 384-well plates (Greiner, Monroe, NC). The following day, plates were washed using an automated plate washer (Tecan, Männedorf, Zürich, Switzerland) with 0.1% BSA and 0.02% Tween-20. Antigen-specific antibody titers were detected with Phycoerythrin (PE)-coupled antibodies against IgG1, IgG2, IgG3, IgG4, IgA1, and IgM (SouthernBiotech, Birmingham, AL). To measure antigen-specific Fc-receptor binding, biotinylated Fc-receptors (FcR2AH, 2B, 3AV, 3B, FcRn, FCAR, FCR3AV – Duke Protein Production facility, C1q – Sigma Aldrich) were coupled to PE to form tetramers and then added to immune-complexed beads to incubate for 1 hour at room temperature while shaking. Fluorescence was detected using an Intellicyt iQue with a 384-well plate handling robot (PAA) and analyzed using Forecyt software by gating on fluorescent bead regions. PE median fluorescence intensity (MFI) was measured as the readout of each antigen-specific antibody measurements. All experiments were performed in duplicate while operators were blinded to study group assignment and all cases and controls were run at the same time to avoid batch effects. The mean value of the duplicate measurements was used for further statistical analysis.

### Ab-Directed *Functional Assays*

Bead-based assays were used to quantify antibody-dependent cellular phagocytosis (ADCP), antibody-dependent neutrophil phagocytosis (ADNP) and antibody-dependent complement deposition (ADCD), as previously described(Ackerman et al., 2018; Ackerman et al., 2011; Fischinger et al., 2019; Karsten et al., 2019; Lu et al., 2016). Yellow (ADNP and ADCP) as well as red (ADCD) fluorescent neutravidin beads (Thermo Fisher) were coupled to biotinylated SARS-CoV-2 S antigens and incubated with diluted plasma (ADCP 1:100, ADNP 1:50, ADCD 1:10) to allow immune complex formation for 2h at 37°C. To assess the ability of sample antibodies to induce monocyte phagocytosis, THP-1s (ATCC) were added to the immune complexes at 1.25E5cells/ml and incubated for 16h at 37°C. For ADNP, primary neutrophils were isolated via negative selection (Stemcell) from whole blood. Isolated neutrophils at a concentration of 50,000 per well were incubated with immune complexes for 1h incubation at 37°C. Neutrophils were stained with an anti-CD66b PacBlue detection antibody (Biolegend) and fixed with 4% paraformaldehyde (Alfa Aesar). To measure antibody-dependent deposition of C3, lyophilized guinea pig complement (Cedarlane) was reconstituted according to manufacturer’s instructions and diluted in gelatin veronal buffer with calcium and magnesium (GBV++) (Boston BioProducts) and mixed with immune complexes. After a 20-minute incubation at 37°C, C3 was detected with an anti-C3 fluorescein-conjugated goat IgG fraction detection antibody (Mpbio). Antibody-dependent NK (ADNK) cell activity was measured via an ELISA-based assay, as described previously (Chung et al., 2015). Briefly, plates were coated with 3mg/mL of antigen (SARS-CoV-2 S) and blocked overnight at 4°C. NK cells were isolated the day of the assay with negative selection (RosetteSep – Stem Cell Technologies) from healthy buffy coats (MGH blood donor center). Diluted plasma samples were added to the antigen-coated plates (1:25 dilution) and incubated for 2h at 37°C. NK cells were mixed with a staining cocktail containing anti-CD107a BV605 antibody (Biolegand), Golgi stop (BD Biosciences) and Brefeldin A (BFA, Sigma Aldrich). 2.5E5 cells/ml were added per well to the immune complexes and incubated for 5h at 37°C. Next, cells were fixed (Perm A, Invitrogen) and stained for surface markers with anti-CD3 APC-Cy7 (BioLegend) and anti-CD56 PE-Cy7 (BD Biosciences). Subsequently, cells were permeabilized using Perm B (Invitrogen) and intracellularly stained with an anti-MIP-1ß-BV421 (BD Biosciences) and IFNγ-PE (BioLegend) antibodies.

All assays were acquired via flow cytometry with iQue (Intellicyt) and an S-Lab 384-well plate handling robot (PAA). For ADCP, events were gated on singlets and bead-positive cells. For ADNP, neutrophils were defined as CD66b positive events followed by gating on bead-positive neutrophils. A phagocytosis score was calculated for ADCP and ADNP as (percentage of bead-positive cells) x (MFI of bead-positive cells) divided by 10000. For ADCD, complement deposition was reported as the median fluorescence intensity of C3 deposition on Spike-coupled beads. For ADNK, NK cells were defined as CD3- and CD56+ events. NK cell activation was quantified as the percentage of NK cells positive for the degranulation marker CD107a (Alter et al., 2004) and for two markers of NK cell activation, MIP-1ß, and IFNγ(Colucci et al., 2003). In the text, we referred to these readouts as CD107aNK, MIP-1ßNK, and IFNγNK.

### Data Pre-processing

Duplicate measurements of antibody isotypes, subclasses, FcR-binding levels and ADCD measurements were averaged for each sample and then log10 transformed. Duplicate measurements of ADNK, ADCP, and ADNP were averaged for each sample. In order to remove antibody features with low magnitude signals, we used the variation in the control samples as a cut off. More specifically, we removed antibody features whose maximum signal in the CCP recipients was less than four standard deviations over the negative control PBS wells (Mean PBS + 4x PBS standard deviation).

### Visualization

The heatmaps were created with the function pheatmap in R package ‘pheatmap’ (version 1.0.12). To eliminate the effect of extreme values and visualize the predominant differences clearly, the color ranges were equally divided into 100 intervals by the quantile range of the percentage of adjusted values across all the measurements. The UMAP visualization was performed on principal components whose cumulative explained variance is larger than 90% by umap function in R package ‘umap’ (version 0.2.7.0) with fine-tuning parameters (neighbor = 8, min.dist = 0.1), and visualized by ggplot function in R package ggplot2 (version 3.3.5)

### Polar Plots

Polar plots were used to visualize the mean percentile of groups in Figure 2C, 3C, 5A. Percentile rank scores were determined for each feature across all considered samples using the function ‘percent_rank’ of the R package ‘dplyr’ (version 1.0.5).

Polar plots for Supplemental **Figure 1C** were used to visualize the S-specific individual antibody profile of CCP-treated and control participants over the course of the clinical trial. Each feature across the respective populations was scaled by min-max normalization.

### Multivariate Models

#### 1. Four Parameters Logistic Regression Model

The details of four parameters logistic regression model were explained in our previous paper.(Zohar et al., 2020) Briefly, all the measurements were normalized to make sure the minimal values across groups were zero and the maximum values were one. To determine the difference of fitted models in each antibody feature involved in the control and treatment groups, the dynamics along the days since symptom onset were described in each group (CCP-treated and control) at the population level using a four-parameter logistic growth curve. Furthermore, to detect differences explained by different explicit parameters between control and CCP-treatment group, we built two paired models simultaneously, allowing for combinations of parameters to differ between the two groups, while the others are shared between the groups. 16 models controlled by the combination of four parameters were evaluated by Akaike Information Criterion (AIC) to balance the model fitness and model complexity. Finally, the best model was picked with the lowest AIC values. Additionally, to analyze the overall difference in parameters across the groups (Fig. 2e), the maximum likelihood estimates for all the models were combined by weighing the contribution of individual models by the Akaike weight.

#### 2. Regression with Clinical Severity Score

The regression model in Figure 2H was trained to associate Clinical Severity Score with top 30 features suggested by the four parameters logistic regression model with a minimal set of features. First, we applied the least absolute shrinkage and selection operator (LASSO) feature selection algorithm to extract significant features. Here, we run the LASSO feature selection 10 times on the whole dataset and picked the set of features, whose occurrences are more than seven times. The details were implemented in the function ‘select_lasso’ in systemseRology R package (v.1.0). Then, a partial least square regression model was trained using the Lasso-selected features. Model performance was evaluated by five-fold cross-validation and the negative models were constructed from permuted labels with multiple iterations. The permuted control models were generated 20 times by shuffling labels randomly for each iteration. Coefficient of determination, denoted as R^2^ was used to evaluate the regression performance. For PLS-R, we use the ‘opls’ function in R package ‘ropls’ (v.1.22.0) for regression and functions in systemseRology R package for the purpose of visualization. To further investigate the predicted performance of clinical severity score using the features selected by LASSO, the whole samples were divided into two groups (higher severity and lower severity) using the threshold 20. Then, the predicted clinical severity scores for all the involved samples were predicted using five-folds cross-validation for 100 repetitions. After that, the averaged ROC curve with the roc curve from each repetition were visualized by roc function in R package pROC (version 1.18.0) as depicted in Figure 2K.

#### 3. Network Analysis

The correlation networks were used to visualize the additional immune measurements significantly associated with the LASSO-selected features, indicating enhanced insights of biological mechanisms. The measurements that were significantly (p value < 0.05) correlated to the selected features after a Benjamini-Hochberg correction were defined as co-correlates. Significant spearman correlations above a threshold of |r| > 0.7 were visualized within the networks. In detail, the spearman correlation coefficients were calculated using ‘rcorr’ function in ‘Hmisc’ package (v4.4.2) and the p values were corrected b “Benjamini-Hochberg’ correction in ‘stats’ package (v.4.0.3). For the purpose of visualization, the correlation networks were visualized using ‘ggraph’ (v.2.0.4) and ‘igraph’ (v.1.2.6) packages.

#### 4. Mixed Linear Model

We used two nested mixed linear models (null and full model) without/with treatment group information to assess the significance of the association between measured antibody levels and treatment groups while controlling for potential confounding clinical characteristics. We fit two mixed linear models and estimated the improvement in model fit by likelihood ratio testing to identify the associated measurements for participant timepoints from the first two weeks of the trial (D1, 3, 8, and 15).

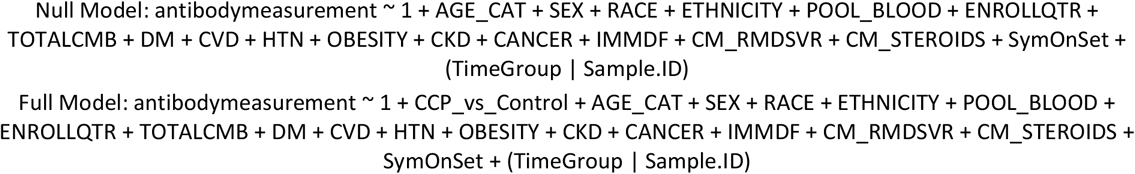

Likelihood ratio test:

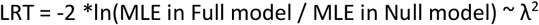

Here, the subject clinical information includes age at enrollment category, gender, race, ethnicity, blood type and enrollment period (May-Jun 2020, July-Aug 2020, Sep-Oct 2020, Nov-Jan 2021). In addition, we included the total number of COVID-19 disease severity modulating comorbidities (TOTALCMB), diabetes (type1, type2) (DM), obesity (OBESITY), hypertension (HTN), cardiovascular disease (CVD), pulmonary disease, chronic kidney disease (CKD), chronic liver disease, cancer (CANCER) and immune deficiency (IMMDF). Additionally, the model also included whether the patients were treated with the drug Remdesivir (CM_RMDSVR) or Steroids (CM_STEROIDS) at baseline and how long they had been symptomatic from COVID-19 (SymOnSet). The R package “lme4” was used to fit the mixed linear model to each measurement and test for difference in antibody features depending on whether a patient received CCP or not.

The P value from the likelihood ratio test and t value (normalized coefficients) associated with the variable represented two arms of the clinical trial, CP_vs_Control in full model, were visualized in a volcano plot using the ggplot function in R package ‘ggplot2’ (Version 3.3.5).

#### 5. Linear Model to identify the percentage of explained variance

We built a linear model to identify the association of Nucleocapsid-related antibody features with clinical outcomes in CCP-treated and control individuals. The linear model used the same clinical characteristics involved in our previous models and N-related measurements from the second week (Day 8 and Day 15 measurements) of the clinical trial to predict the clinical outcomes as measured by the CSC. The details of the linear model are shown as follows:

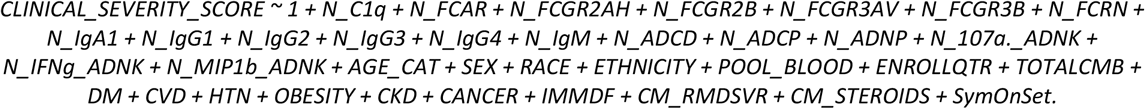

Then, the percentage of explained variance in CSC attributed to each antibody feature was calculated by Sum of Square in analysis of variance (ANOVA).

#### 6. Unsupervised Clustering to identifying the patterns in Day0

Using all the measurements or the selected subset of measurements, the spearman correlation coefficients across the measurements were calculated to represent the sample-sample similarities. Then, we applied community detection method to the similarity matrix to identify the groups with more homogeneous immune profiles. First, we made a K-nearest neighbor graph based on similarity distance. Secondly, we calculated the adjacent matrix and identified the communities using the R package ‘igraph’. Here, the parameter K was searched exhaustively from low number (2) to high number, in which all the samples were grouped into one cluster. The number of clusters was selected with the largest averaged silhouette values across all the clustering results.

To identify the measurements distinguished cluster four from the other clusters (Cluster 1,2, 3), we compared the samples inside the cluster four and those outside it using the wilcox-rank test implemented in the wilcoxauc function, in the presto R package. This process was repeated only with the 14 measurements that made up the CCP benefit signature to create Cluster A and B. The same clustering procedure described above was followed to determine cluster A and B.

To evaluate whether antibody levels or function was a more important determinant of response to CCP therapy, we created three linear models based on following measurements that most distinguished Cluster 4 from Cluster 1, 2, 3: 1) the 14 antibody titers, 2) the 14 antibody functions w/o IgG1 normalization, and 3) 14 antibody functions w/ IgG1 normalization. These models were fitted and the percentage of explained variance was determined with the R package “lme4”.

#### 7. Discriminant Analysis in Day60

The log2 Fold change of the average value per each measurement was calculated between CCP treatment arm and control arm. The p values were estimated by the permutation test through shuffling the arm labels. In detail, we randomly shuffled the arm labels and recalculated the log2 Fold change of the mean values in the two groups for 1000 times and then the P values were estimated by the rank of the actual Fold change among the values of shuffled fold changes.

Then, the classification models were trained to distinguish control and treatment groups with a minimum set of measurements. We first applied LASSO feature selection and then trained partial least squares discriminant analysis (PLS-DA) classifier on the selected features as described above. The model performance was evaluated by five-fold cross-validation. Finally, the network analysis was used to investigate the correlated features between selected features and non-selected features. The significant spearman correlation above the threshold of |r| > 0.3, were visualized within the networks.

### Fc Glycan Analysis

Capillary electrophoresis was conducted as previously described.(Mahan et al., 2015) Briefly, recombinant S was biotinylated and coupled to 1 μm neutravidin-coated magnetic beads; 5 μg of protein was coupled to 50 μL of beads for each sample. Heat-inactivated sample (100 μL) was incubated with 50 μL of un-coupled magnetic beads to clear non-specific bead binding for 30 minutes. Pre-cleared plasma was incubated with 50 μL of protein-coupled beads and incubated for one hour at 37°C, were washed, and the captured antibody Fc was cleaved off by incubating with 1 μL of IDEZ at 37°C for 1 hour. The isolated <u>Fc fragments</u> were deglycosylated and the freed glycans were fluorescently labeled and purified using a GlycanAssure APTS Kit according to the manufacturer’s instructions. Glycans were analyzed by capillary electrophoresis on 3500xL genetic analyzer (Applied Biosystems). Samples were run with N-glycan fucosyl, afucosyl, bisecting and mannose N-glycan libraries to enable identification of twenty-two discrete glycan species. Glycan profiles of each labeled and purified participant sample was measured with technical duplicate The relative frequencies of each glycan peak were plotted as a percentage of total glycans, calculated using GlycanAssure software.

## Data Availability

All data produced in the present study are available upon reasonable request to the authors.

## Study Approval

The clinical cohort described in Bar et al.(Bar et al., 2021) was approved by the University of Pennsylvania institutional review board and registered at ClinicalTrials.gov with number NCT04397757. All participants provided informed consent prior to participation in the study. Secondary Use of patient samples and clinical samples was approved by the Mass General Brigham Institutional Review Board.

## Data Availability

The dataset generated during and/or analyzed during the current study have been made available in the Supplementary material.

## Code Availability

Custom code was used in this manuscript and has been made available at Zenodo.org under the record number 6110200. The R packages used for data analysis are described in more detail in the Methods section and more information is available upon request.

## Acknowledgements

We thank Nancy Zimmerman, Mark and Lisa Schwartz, an anonymous donor (financial support), Terry and Susan Ragon, and the SAMANA Kay MGH Research Scholars award for their support. We acknowledge support from the Ragon Institute of MGH, MIT and Harvard, the Massachusetts Consortium on Pathogen Readiness (MassCPR), the NIH (3R37AI080289-11S1, R01AI146785, U19AI42790-01, U19AI135995-02, U19AI42790-01, 1U01CA260476 – 01, CIVIC75N93019C00052, T32 AI007061), the Gates Foundation, the Global Health Vaccine Accelerator Platform funding (OPP1146996 and INV-001650), and the Musk Foundation.

## Authorship Contributions

J.D.H., H.Y., L. P., B.J., D.L., K.J.B., and G.A. conceived of the idea. K.J.B., P.A.S, G.H. C., P.T., D.S., and I.F. designed, conducted, analyzed the results of the randomized clinical trial.

J.D.H. and G.A. designed the experiments. J.D.H., J.S.B, Y.Z., H.C., J.K., R.M., and S.S. performed the antibody profiling experiments. J.D.H., C.W., D. L, and G.A. analyzed the data. And J.D.H.,C.W.,K.J.B, D.L., and G.A. wrote the paper with input from all authors.

---

**Supplementary Figure 1:**
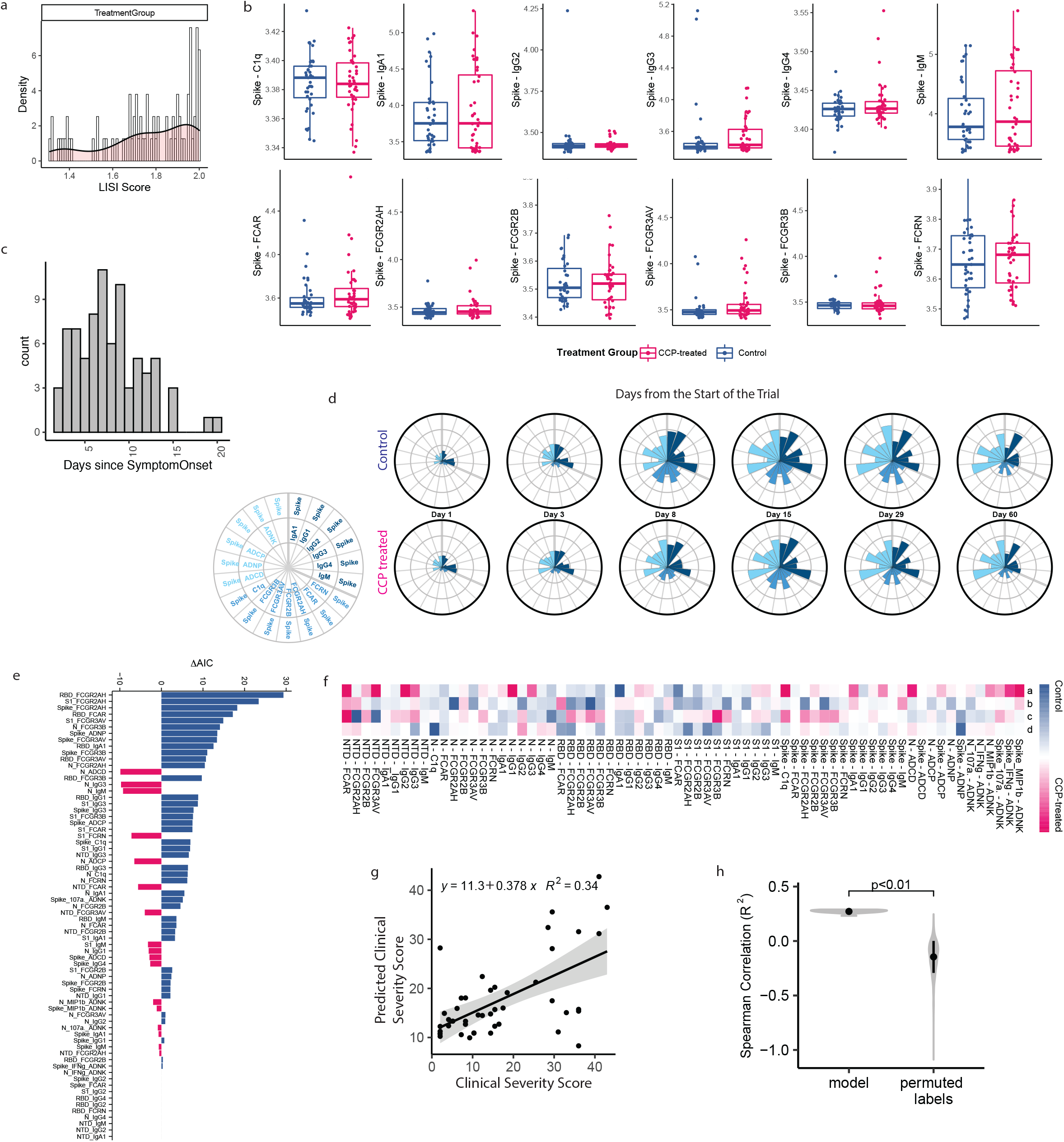
Supplement to Figure 2. (A) This histogram shows the distribution of Local Inverse Simpson’s Index (LISI) scores for treatment group in UMAP visualization of all patient samples profiled in this paper shown in Fig. 2A. LISI measures the degree of mixing in an embedding ranging from 1 to the number of categories (here, will be two), where larger LISI scores indicate less separation and more homogenous mixing. (B) Univariate box plots of Day 1 (pre-existing) Spike specific antibody features of CCP-treated and control patients. Each box represents the median (central line) and IQR (25% and 75% percentiles) and the two whiskers represent 1.5 * IQR. (C) The histogram shows the distribution of days of symptomatic COVID-19 prior to enrollment for all participants. (D) The polar plots depict the min-max proportion of each Spike-specific antibody features at each sample day across the control (top) and CCP-treated (bottom) participants. Antibody features are colored by type of feature with ab-directed functions in light blue, Fc receptor binding in medium blue, and Ab titer in dark blue. (E) The bar plot depicts the delta-AIC of the best model compared with the model without differences of the four parameters. An abbreviated form of just the top 30 features is included in Figure 2F. The higher the delta-AIC, the better the model can explain the trajectory difference. The sign of delta-AIC represents the AUC difference between the CCP-treated and control curves, thereby showing whether the antibody feature is enriched in the CCP-treated model (negative) or the control model (positive). The bars are colored according to whether the feature is enriched in the CCP-treated model (pink) or control model (blue). (F) This heatmap shows the Akaike Information Criterion (AIC) weighted average parameter differences between the two groups. Each column shows a parameter, which is normalized across the features. The color intensity indicates whether the parameter is higher in the CCP-treated (blue) or control (orange) model. An abbreviated form of this heatmap is included in Figure 2E. (G) To validate the PLS-R model in Figure 2H of Clinical Severity based on the Top 30 features, we calculated the correlation between the predicted clinical severity score from five-folder cross-validation and the actual clinical severity score. (H) To further demonstrate the performance and robustness of the PLS-R model, we compared the cross-validation result with a randomly permuted dataset. The violin plots show the distribution of repeated five-fold cross validation R^2^ values using the actual data (model) or shuffled labels (permuted labels). Black dot indicates the median values and whiskers represent the one standard variance.

**Supplementary Figure 2:**
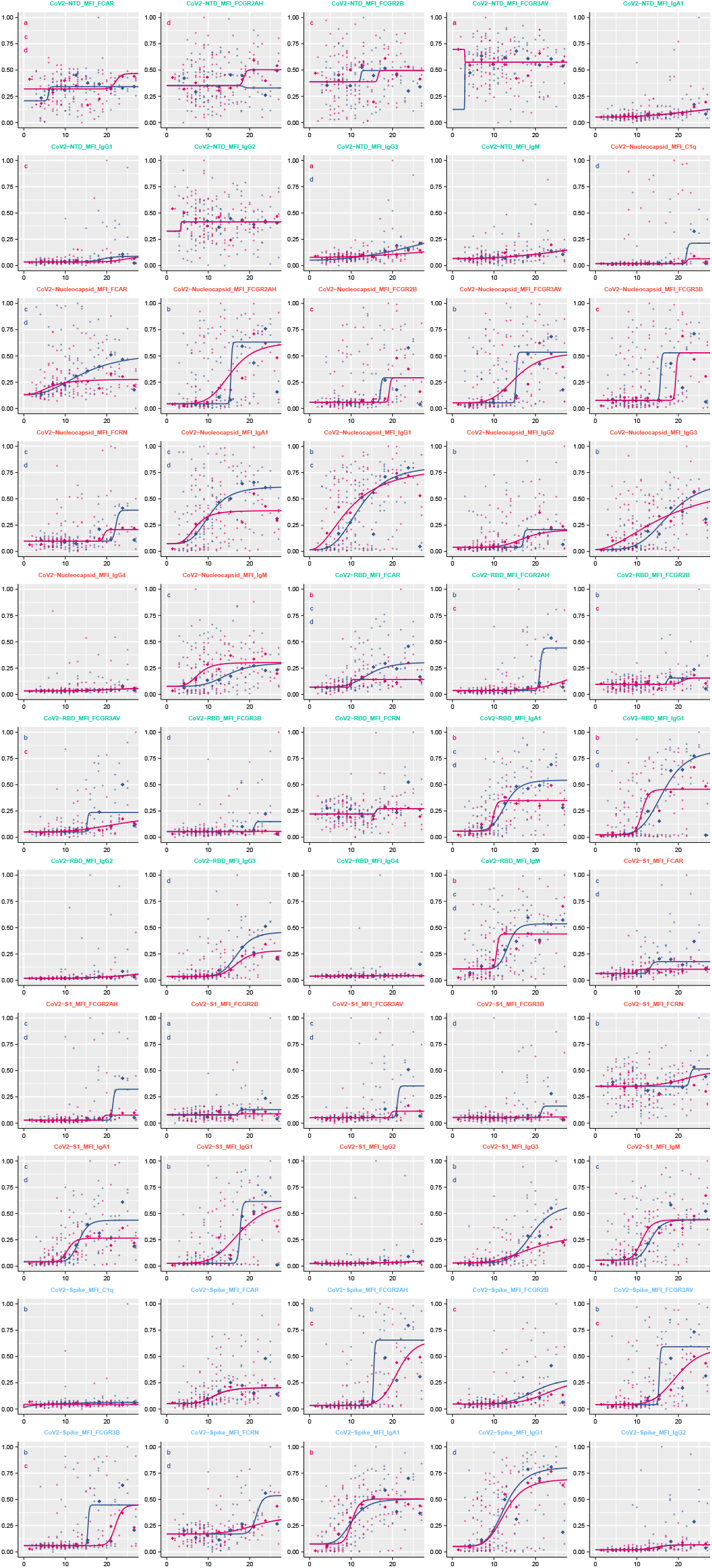
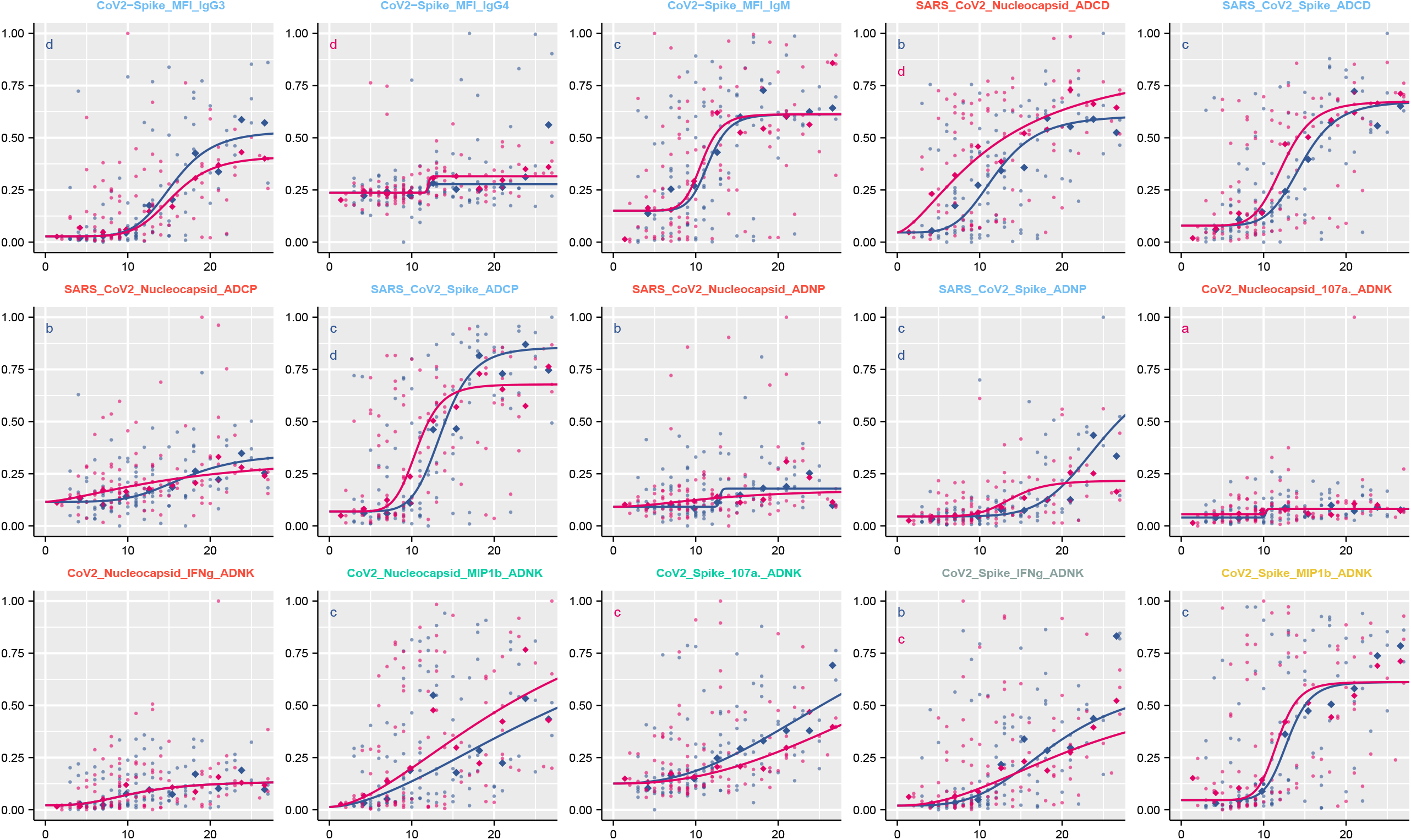
Fitted temporal evolutionary curves of antibody features by our regression model depicted in Figure 2 D-G. For each antibody feature, the optimal model fit is shown for each group. Dots indicate individual patients, diamonds indicate the binned median, the curves indicate the optimal fitted model, and the color shows the group. The parameters which are different for the displayed model are indicated in the top left corner and color-coded according to the group for which the parameter is higher.

**Supplementary Figure 3:**
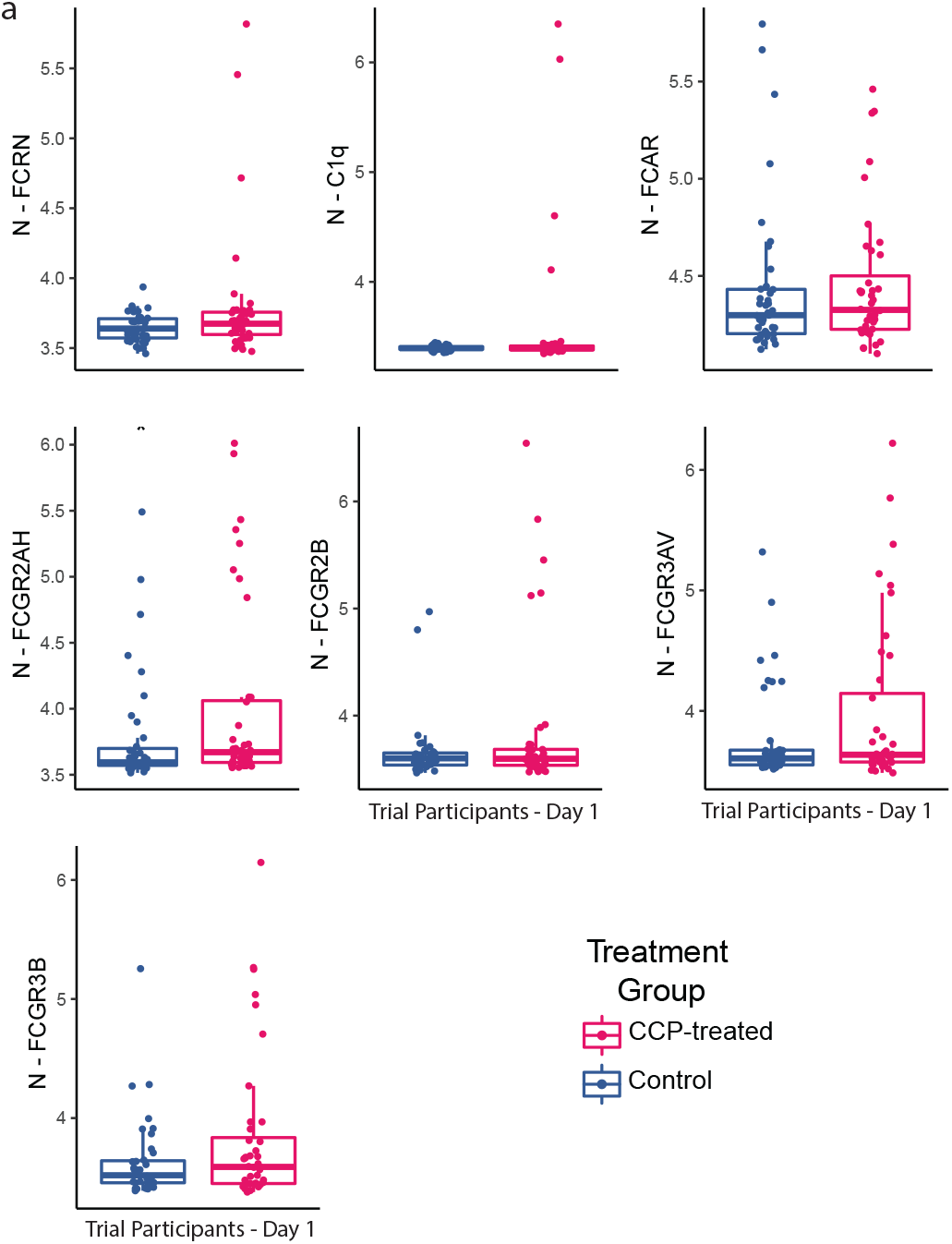
(A) Univariate box plots on Nucleocapsid specific measurements of CCP-treated and Control patients on Day 1(Pre-existing). Each box represents the median (central line) and IQR (25% and 75% percentiles) and the two whiskers represent 1.5 * IQR.

**Supplementary Figure 4:**
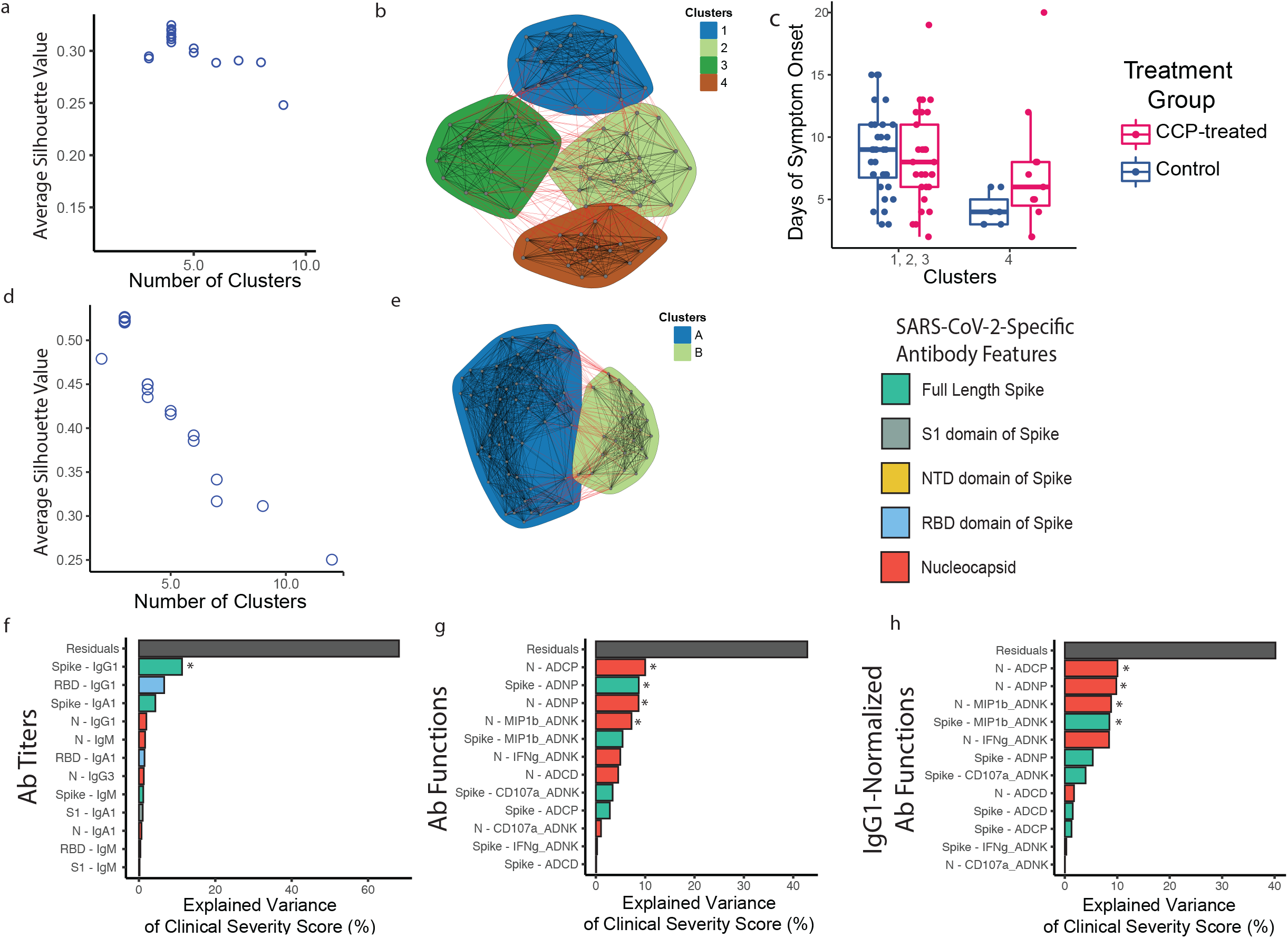
Supplement to Figure4. (A, B) Average Silhouette values in Spearman correlation-based community detection algorithm were used to select the number of clusters. (A) Point plot of the average silhouette values for 1 through 10 clusters using the community detection algorithm to separate participants by their pre-existing SARS-CoV-2 antibody profile. (B) Network plot of individual participants arranged into the four clusters selected by Silhouette analysis. Each dot represents each sample, and each colored region indicates each cluster defined by community detection algorithm. Clusters are indicated by color. (C) Boxplot of Day of Symptom Onset prior to enrollment in the clinical trial of CCP-treated and Control patients in Cluster 4 and Cluster 1, 2, 3. A two-sided Wilcoxon test was performed to compare age between treatment arms. (D) Participants were re-clustered by their CCP benefit signature, identified in Figure 4D, and we used silhouette analysis to choose the number of clusters. This point plot shows the calculated average silhouette values for 1 through 15 clusters using the community detection algorithm to separate participants by their CCP benefit signature identified in Figure 4D. (E) Network plot of individual participants arranged into two clusters. Each dot represents each sample, and each colored region indicates each cluster defined by community detection algorithm. Clusters are indicated by color. (F-H) Three separate linear regression models were used to assess which type of pre-existing antibody features best predicted clinical severity in CCP-treated individuals. The bar plots show the percentage of explained variance (%) by all 12 antibody titers (F), all 12 antibody functions (G), or all 12 IgG1 titer-corrected antibody functions (H) in the three separate models. The bars are colored by the SARS-CoV-2 antigen for which they correspond, and the asterisks represents Ab features with a statistically significant relationship with clinical severity score. For the boxplots in C, each box represents the median (central line) and IQR (25% and 75% percentiles) and the two whiskers represent 1.5 * IQR. * represents a p-value < 0.05.

**Supplementary Figure 5:**
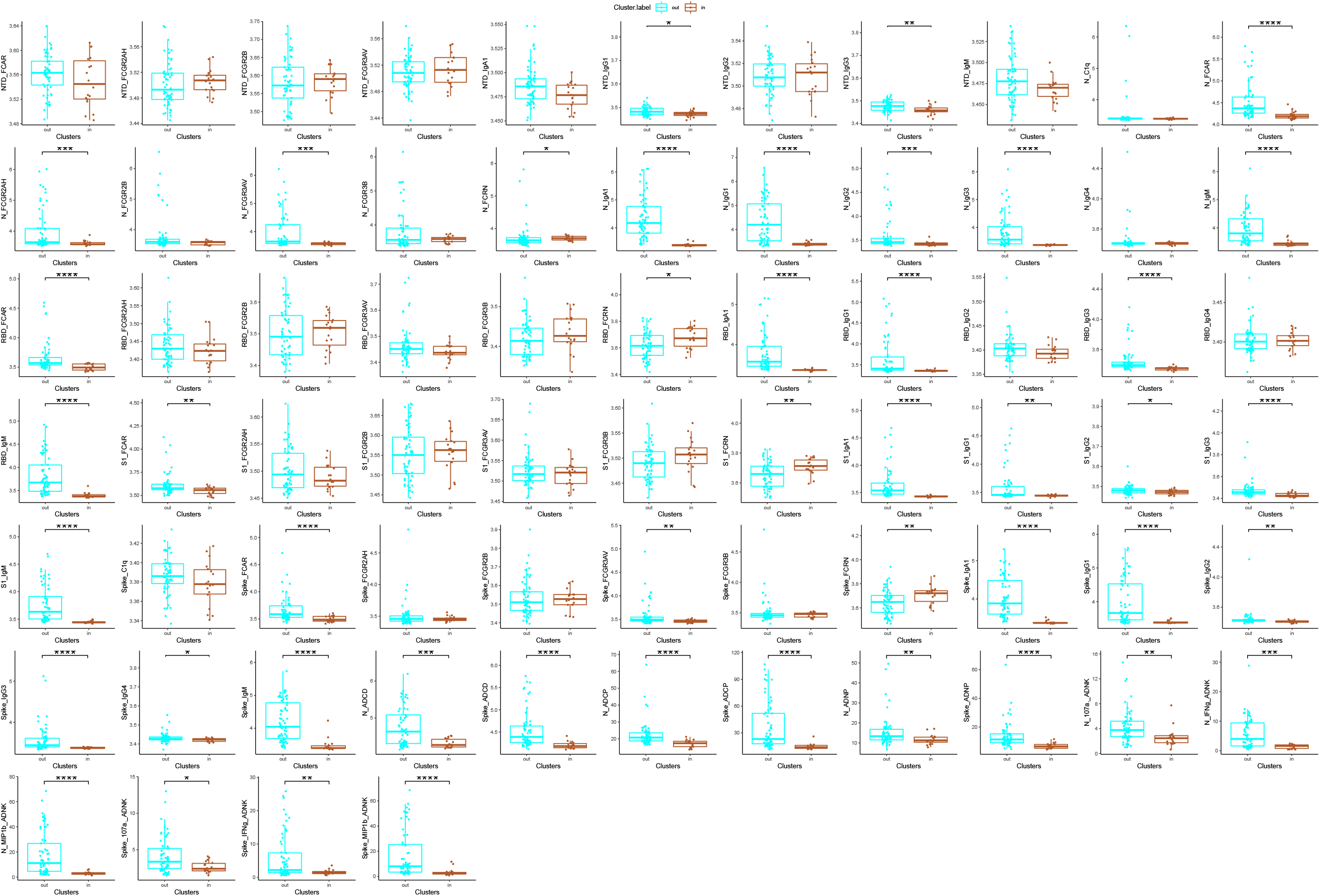
Univariate box plots for all antigen-specific antibody measurements of CCP-treated and control patients in Cluster 4 (in) and Cluster 1,2,3 (out), described in Figure 4. Each box represents the median (central line) and IQR (25% and 75% percentiles) and the two whiskers represent 1.5 * IQR. The difference antigen-specific antibody features between CCP-treated and control patients was tested by a two-sided Wilcoxon test. * represents a p-value < 0.05. ** represents a p-value <0.01. *** represents a p-value <0.001. **** represents a p-value < 0.0001.

**Supplementary Figure 6:**
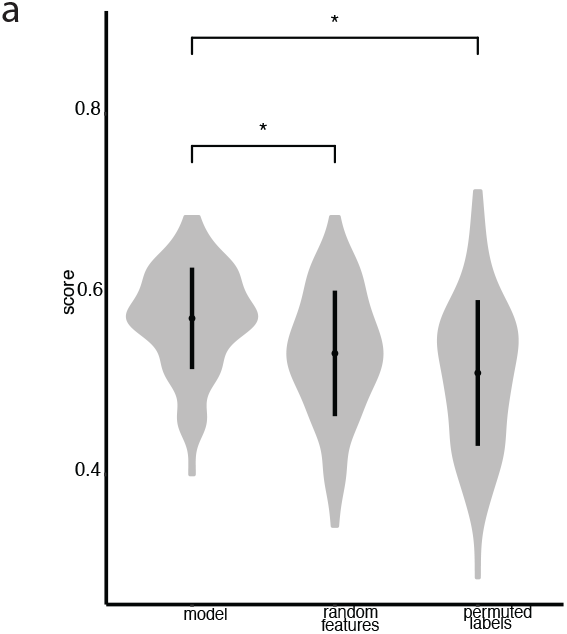
(A) The PLS-DA model in Figure 5 C, D was found to have an accuracy of 63.8% and under a five-fold cross-validation framework run 100 times and preformed significantly better than both negative models. The violin plots show the distribution of repeated cross validation R^2^ values using the actual data (model), randomly selected antibody features (random features) or shuffled participant labels (permuted labels). The whiskers represent the one standard variance and * represents a p-value < 0.0001 by two-tailed Mann Whitey U testing.

**Supplementary Table 1:** Table of the Clinical Characteristics of Pre-existing Ab Cluster 4 vs. Clusters 1,2,3

| **Demographics** |  | **Cluster 4 (n=18)** | **Cluster 1,2,3 (n=61)** | **p-value (Fisher's Exact test)** |
| --- | --- | --- | --- | --- |
| Age - Median (IQR) |  | 66.5(32.75) | 62(20) | 0.35 |
| Sex |  |  |  | 0.79 |
|  | Male | 9 | 27 |  |
|  | Female | 9 | 34 |  |
| Race |  |  |  | **0.033** |
|  | African-American | 5 | 37 | . |
|  | Asian | 1 | 3 | . |
|  | Caucasian | 10 | 20 | . |
|  | Unknown | 2 | 1 | . |
| Ethnicity |  |  |  | 0.13 |
|  | Hispanic | 2 | 1 |  |
|  | Non-hispanic | 16 | 60 |  |
| Blood Group |  |  |  | 0.58 |
|  | A | 5 | 23 |  |
|  | B | 1 | 7 |  |
|  | O | 12 | 31 |  |
| **Study Characteristics** |  |  |  |  |
| Enrollment Quarter |  |  |  | 0.021 |
|  | May - June 2020 | 2 | 17 |  |
|  | July-August 2020 | 1 | 18 |  |
|  | September-October 2020 | 4 | 6 |  |
|  | November-January2021 | 11 | 20 |  |
| **COVID-19 Co-morbidities** |  | 18 | 61 |  |
| Cardiovascular disease - no. |  | 9 | 18 | 0.16 |
| Cancer - no. |  | 7 | 14 | 0.23 |
| Obesity - no. |  | 4 | 32 | **0.031** |
| Chronic Kidney Disease - no. |  | 10 | 16 | **0.026** |
| Hypertension - no. |  | 11 | 42 | 0.58 |
| Diabetes - no. |  | 6 | 26 | 0.59 |
| **Treatments** |  |  |  |  |
| Immodulatory Treatment - no. |  | 4 | 7 | 0.26 |
| Remdesivir - no. |  | 14 | 50 | 0.74 |
| Corticosteroids - no. |  | 14 | 52 | 0.48 |

**Supplementary Table 2:** Table of the Clinical Characteristics of Pre-existing Ab Cluster where CCP provides Benefit: Cluster 4 CCP-treated vs. Control Patients

| **Demographics** |  | **CCP-treated (n=7)** | **Control (n=11)** | **(Fisher's Exact test )** |
| --- | --- | --- | --- | --- |
| Age - Median (IQR) |  | 86(14) | 64(30.5) | 0.0184 |
| Sex |  |  |  | 1 |
|  | Male | 3 | 6 |  |
|  | Female | 4 | 5 |  |
| Race |  |  |  | **0.25** |
|  | African-American | 1 | 4 | . |
|  | Asian | 0 | 1 | . |
|  | Caucasian | 6 | 4 | . |
|  | Unknown | 0 | 2 | . |
| Ethnicity |  |  |  | 1 |
|  | Hispanic | 1 | 1 |  |
|  | Non-hispanic | 6 | 10 |  |
| Blood Group |  |  |  | 0.75 |
|  | A | 3 | 2 |  |
|  | B | 0 | 1 |  |
|  | O | 4 | 8 |  |
| **Study Characteristics** |  |  |  |  |
| Enrollment Quarter |  |  |  | 0.88 |
|  | May - June 2020 | 1 | 1 |  |
|  | July-August 2020 | 0 | 1 |  |
|  | September-October 2020 | 1 | 3 |  |
|  | November-January2021 | 5 | 6 |  |
| **COVID-19 Co-morbidities** |  |  |  |  |
| Cardiovascular disease - no. |  | 5 | 4 | 0.33 |
| Cancer - no. |  | 4 | 3 | 0.33 |
| Obesity - no. |  | 1 | 3 | **1** |
| Chronic Kidney Disease - no. |  | 6 | 4 | **0.066** |
| Hypertension - no. |  | 6 | 5 | 0.15 |
| Diabetes - no. |  | 3 | 3 | 0.63 |
| **Treatments** |  |  |  |  |
| Immodulatory Treatment - no. |  | 1 | 3 | 1 |
| Remdesivir - no. |  | 5 | 9 | 1 |
| Corticosteroids - no. |  | 6 | 8 | 1 |

**Supplementary Table 3:** Table of the Clinical Characteristics of CCP Benefit Signature Clustered Groups: Cluster A vs. Cluster B

| **Demographics** |  | **A (n=57)** | **B (n=22)** | **p-value (Fisher's Exact test)** |
| --- | --- | --- | --- | --- |
| Age - Median (IQR) |  | 64(22) | 60.5(20.75) | 0.336 |
| Sex |  |  |  | 0.626 |
|  | Male | 27 | 9 |  |
|  | Female | 30 | 13 |  |
| Race |  |  |  | 0.498 |
|  | African-American | 29 | 13 | . |
|  | Asian | 2 | 2 | . |
|  | Caucasian | 23 | 7 | . |
|  | Unknown | 3 | 0 | . |
| Ethnicity |  |  |  | 1 |
|  | Hispanic | 2 | 1 |  |
|  | Non-hispanic | 55 | 21 |  |
| Blood Group |  |  |  | 0.579 |
|  | A | 22 | 6 |  |
|  | B | 5 | 3 |  |
|  | O | 30 | 13 |  |
| **Study Characteristics** |  |  |  |  |
| Enrollment Quarter |  |  |  | 0.871 |
|  | May - June 2020 | 14 | 5 |  |
|  | July-August 2020 | 15 | 4 |  |
|  | September-October 2020 | 7 | 3 |  |
|  | November-January2021 | 21 | 10 |  |
| **COVID-19 Co-morbidities** |  |  |  |  |
| Cardiovascular disease - no. |  | 19 | 8 | 0.797 |
| Cancer - no. |  | 14 | 7 | 0.574 |
| Obesity - no. |  | 25 | 11 | 0.801 |
| Chronic Kidney Disease - no. |  | 20 | 6 | 0.599 |
| Hypertension - no. |  | 39 | 14 | 0.791 |
| Diabetes - no. |  | 22 | 10 | 0.616 |
| **Treatments** |  |  |  |  |
| Immodulatory Treatment - no. |  | 9 | 2 | 0.718 |
| Remdesivir - no. |  | 45 | 19 | 0.539 |
| Corticosteroids - no. |  | 47 | 19 | 1 |

**Supplementary Table 4:** Table of Clinical Characteristics of CCP Benefit Signature Clustered Group where CCP provides benefit: Cluster A CCP-treated vs. Control

| **Demographics** |  | **Control (n=29)** | **CCP-treated (n=28)** | **p-value (Fisher's Exact test)** |
| --- | --- | --- | --- | --- |
| Age - Median (IQR) |  | 62(19) | 65(31.75) | 0.615 |
| Sex |  |  |  | 0.189 |
|  | Male | 11 | 16 |  |
|  | Female | 18 | 12 |  |
| Race |  |  |  | 0.611 |
|  | African-American | 16 | 13 | . |
|  | Asian | 0 | 2 | . |
|  | Caucasian | 12 | 11 | . |
|  | Unknown | 1 | 2 | . |
| Ethnicity |  |  |  | 1 |
|  | Hispanic | 1 | 1 |  |
|  | Non-hispanic | 28 | 27 |  |
| Blood Group |  |  |  | 1 |
|  | A | 11 | 11 |  |
|  | B | 3 | 2 |  |
|  | O | 15 | 15 |  |
| **Study Characteristics** |  |  |  |  |
| Enrollment Quarter |  |  |  | 0.946 |
|  | May - June 2020 | 8 | 6 | . |
|  | July-August 2020 | 7 | 7 | . |
|  | September-October 2020 | 4 | 3 | . |
|  | November-January2021 | 10 | 11 | . |
| **COVID-19 Co-morbidities** |  |  |  |  |
| Cardiovascular disease - no. |  | 7 | 12 | 0.167 |
| Cancer - no. |  | 8 | 6 | 0.76 |
| Obesity - no. |  | 15 | 10 | 0.289 |
| Chronic Kidney Disease - no. |  | 12 | 8 | 0.408 |
| Hypertension - no. |  | 23 | 16 | 0.092 |
| Diabetes - no. |  | 14 | 8 | 0.175 |
| **Treatments** |  |  |  |  |
| Immodulatory Treatment - no. |  | 5 | 4 | 1 |
| Remdesivir - no. |  | 23 | 22 | 1 |
| Corticosteroids - no. |  | 26 | 21 | 0.179 |

